# Many Oil Wells, One Evil: Potentially toxic metals concentration, seasonal variation and Human Health Risk Assessment in Drinking Water Quality in Ebocha-Obrikom Oil and Gas Area of Rivers State, Nigeria

**DOI:** 10.1101/2021.11.06.21266005

**Authors:** Olalekan Morufu Raimi, Olawale Henry Sawyerr, Clinton Ifeanyichukwu Ezekwe, Gabriel Salako

## Abstract

**Background:** Oil and natural gas extraction have produced environmental pollution at levels that affect reproductive health of indigenous populations. Accordingly, polluted drinking water from physical, chemical and heavy metals can result in serious health problems, like anemia, kidney failure, immunosuppression, neurological impairments, gastrointestinal as well as respiratory irritation, skeletal system abnormalities, liver inflammation, liver cancer, cardiovascular diseases after chronic exposure and other cancer diseases with negative health effects. These diseases types remain associated to high amounts of heavy metal elements such as lead, chromium, zinc, copper, cadmium, manganese as well as nickel etc.

**Objectives:** Compare differences in water quality parameters in the study area (determine the level of pollutions in the different sites).

**Methodology:** The investigation made use of standard analytical procedures. All sampling, conservation, transportation and analysis followed standard procedures described in APHA (2012). To prevent degradation of the organic substances, all obtained samples were transferred to the laboratory, while keeping in an icebox.

**Results:** Result shows that during wet season, the mean values obtained for water quality parameters were significantly lower in site 9 compared with that obtained in other sites (p<0.05) with the exemptions of temperature, DO, BOD, COD, acidity, TH, TDS, K, Mg, Zn, Mn, Cd, Pb, Cu, Cr, NH_3_, NO_2_, NO_3_, Ni though slightly lower in most cases in site 9 were not significantly different (p>0.05) and both alkalinity and SO_4_ which were significantly higher in site **9** than site 1 (p<0.05). Result obtained during dry season reveals that there is no remarkable difference in pH, acidity, Pb and Ni between the nine sites (p>0.05) while other water quality parameters were significantly lower in site 9 than other sites excluding Cl and Mg which were both significantly higher in site 9 than site 8 (p<0.05).

**Conclusion:** To guarantee quality groundwater supply for various purposes in Nigeria’s core Niger Delta region, extra efforts must be taken to fully understand hydrogeochemical features and its suitability. Thus, this study will aid in the development of a quantitative understanding of the effects of diverse causes on groundwater level fluctuations in any aquifer around the world. Also, this analysis reinforces a valuable resource for researchers, activists and public officials seeking to help enhance community awareness, planning and performance. The verdicts would remain a valuable guideline for policymakers, the Ministry of Water Resources and development practitioners, as this highlights the requirement for suitable approaches toward mitigating toxic element of water resources contamination in the core Niger Delta toward safeguarding health of the public from carcinogenic as well as non-carcinogenic risks.

**HIGHLIGHTS:**

- Many tropical countries are suffering from severe groundwater pollution. Governments at all levels are doing little or very little to provide clean and accessible water to citizens, especially in Nigeria’s Niger Delta region.
- This study aims to determine the level of pollutions in the different sites.
- Result depicts that during wet and dry season, the mean values obtained for water quality parameters were significantly lower in site 9 compared with that obtained in other sites.
- Result reveals that groundwater at location 3, 4 and 7 were heavily polluted during wet and dry season. Hence, an alliance is needed to address the rising global health emergency threat caused by groundwater pollution in Nigeria’s core Niger Delta region, which is threatening millions of people. The situation will only get worse and faster unless there is a coordinated response to the problem through a worldwide alliance of organizations capable of bringing meaningful change.
- The disease risk as well as illness to millions of individuals living in close proximity to gas flaring remain a cause for worry in its own right, the gases as well as toxins impact released into the atmosphere through continuously flaring gases has worldwide implications.
- Regarding the environmental and social conditions of the area, gas flaring significantly increases the health hazards, first through releasing dangerous pollutants directly into the atmosphere as well as through pollutants transfer to the food chain.
- Groundwater pollution has a financial cost that runs into billions of Naira, in addition to the human and environmental effect. Thus, there is need for tougher environmental regulations.
- At present, no coordinated action being taken, real change will only occur if governments as well as key stakeholder organizations form a global alliance toward addressing the issue. Starting with a strategy to finance well closure as well as relocation of sites that are most dangerous (location 3, 4 & 7) as soon as possible, as well as providing support through capital and experience is required. Even though the cost will remain substantial, it will provide an opportunity toward investing in the Niger Delta infrastructure as well as economy. Furthermore, the expense of closing the most dangerous groundwater open wells will be a drop in the bucket compared to the cost of the health consequences.

## 1. Introduction

The total worldwide fuel consumption of oil as well as gas accounts for roughly 56%. About 100 million barrels of oil as well as 10 billion cubic metres of natural gas are produced daily by the industry. Thus, from operations of the oil and gas, exploration to manufacturing, storing as well as transporting products, can have a significant environmental impact (Morufu *et al.*, 2021a). Consequentially, it’s possible that it will have an effect on human health, biotic as well as abiotic or cultural heritage sites on a local or regional level (Raimi and Sabinus, 2017a; Raimi *et al.*, 2018; Premoboere and Raimi, 2018; Suleiman *et al.*, 2019; Deinkuro *et al.*, 2021; Morufu *et al.*, 2021c; Olalekan *et al.*, 2021). While, the area plays host to over 100 oil wells, 9 gas flaring sites, an oil and gas gathering centre, a gas plant, a gas to power plants and several hundreds of kilometers of oil and gas pipelines all located within an area of less than 250 km^2^ (Morufu and Clinton, 2017; Raimi and Sabinus, 2017b; Olalekan *et al.*, 2018b; Olalekan *et al.*, 2020a; Morufu *et al.*, 2021a). Niger Delta region has a difficult terrain, and as such all kinds of environmental pollution is quite high (Olalekan *et al.*, 2018b; Premoboere and Raimi, 2018; Olalekan *et al.*, 2020a). Water bodies in the Niger Delta have been heavily polluted due to recurring incidence of oil spillage (Olalekan *et al.*, 2018b; Premoboere and Raimi, 2018; Olalekan *et al.*, 2020a). Most micro-populations following several large-scale oil spillages have generally affected aquatic resources (Okoyen *et al.*, 2020; Olalekan *et al.*, 2021). As a result, freshwater resources contamination has come to be an issue of worldwide concern. It has been put forward that it is the largest cause of fatalities as well as diseases, with over 14,000 individual’s deaths daily (West, 2006; Raimi *et al.*, 2017; Olalekan *et al.*, 2018a; Raimi *et al.*, 2019a; Olalekan *et al.*, 2019a; Raimi *et al.*, 2019d; Gift and Olalekan, 2020; Gift *et al.*, 2020; Afolabi and Raimi, 2021). In spite of the about US$600 billion generated from oil revenue between 1958 and 2007, Nigeria has little or nothing to show in terms of social amenities and infrastructural provision (Watts, 2008; Premoboere and Raimi, 2018; Deinkuro *et al.*, 2021). Groundwater is rapidly being exploited for drinking-water supplies, according to studies, partly due to dwindling water resources scarcity as well as because groundwater is valued as a source of “clean” drinking water (Winkel *et al.*, 2008; Amini *et al.*, 2008a; Morufu and Clinton, 2017; Raimi and Sabinus, 2017b; Olalekan *et al.*, 2018b; Olalekan *et al.*, 2020a). Ground water accounts for around half (50%) of the potable water supply in Nigeria. Approximately 80 percent of homes in Niger Delta rely on this domestic source of water supply. Concern about groundwater supply safety have focused on pollution caused by human activities, with contamination from natural origins receiving less attention. The absence of safe water is linked to this, which exacerbates health problems and has a negative impact on productivity. Only around a quarter (24%) of the indigenous population as well as half of the urban population in the Niger Delta have access to drinkable water, according to the UNDP (2006). This is in accordance with the findings of a Bayelsa State Micro Credit Administration Agency poverty baseline survey, which indicated that only a tiny percentage of the indigenous populace has access to drinkable water (Olalekan *et al.*, 2020a). However, several studies (e.g., Raimi and Sabinus, 2017b; Morufu and Clinton, 2017; Olalekan *et al.*, 2018b; Olalekan *et al.*, 2019a; Raimi *et al.*, 2019a; Olalekan *et al.*, 2020a) have shown and document (scientifically) that increasing presence of geogenic contaminants in the Niger Delta can have serious health effects as well as wellbeing on the indigenous population, thus leading to both environmental and community concerns, resulting in oil and gas companies’ prohibition in some locations. The volume as well as frequency of hydrocarbon spills larger than one barrel (bbl) that reach the environment remain reported as normal industry practice. For reference, 1 barrel equals 42 gallons or 159 litres in the United States. As a results, figure 1 below illustrates some of the most important sustainability challenges for the extractive industry. It likewise demonstrates how social, economic as well as environmental factors are all inter-connected in nature. This covers a wide variety of positive as well as negative impacts that companies may face while addressing sustainability risks as well as opportunities.

**Figure 1:**
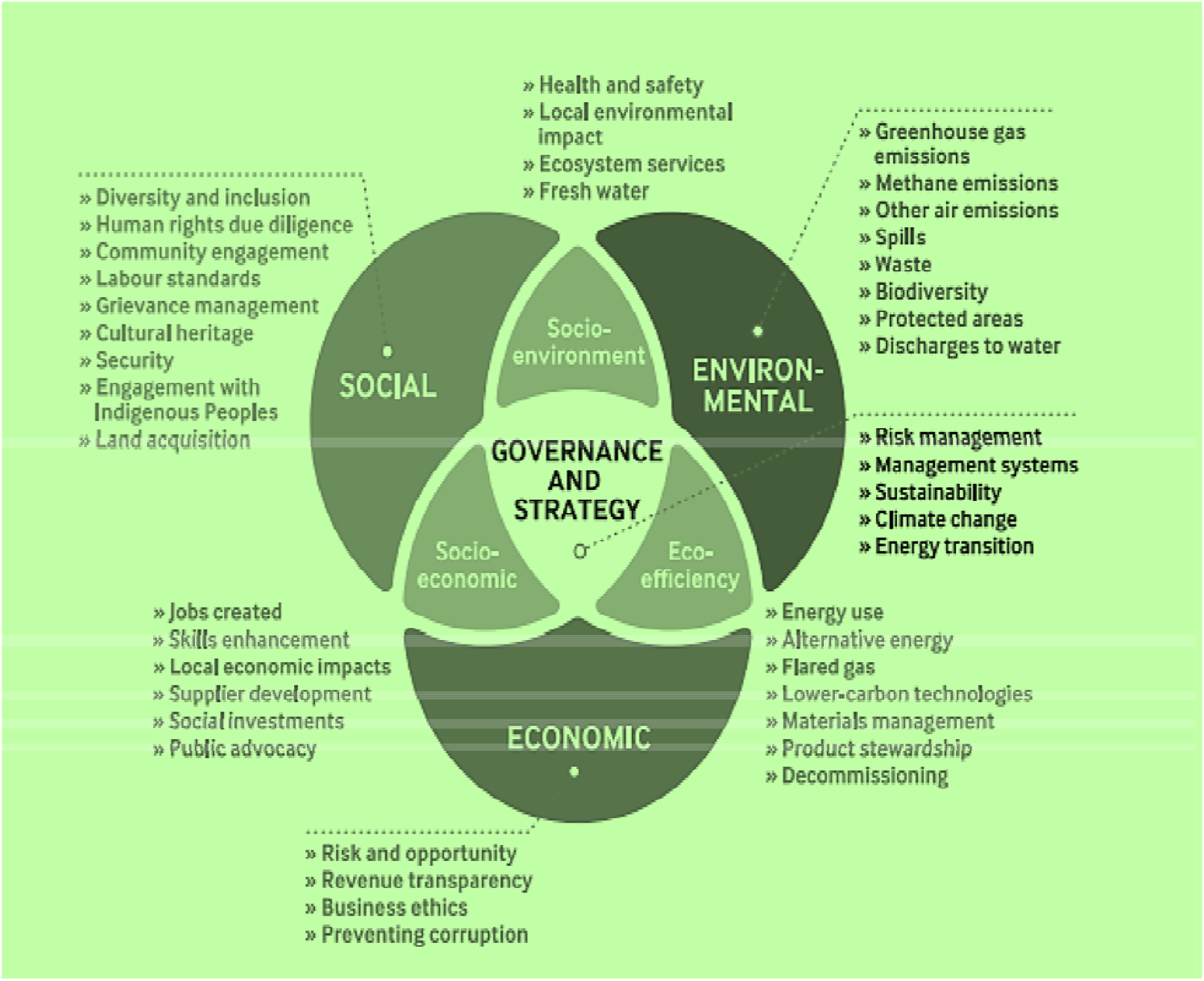
Sustainability issues in the oil and gas industry. Adapted from IPIEA (2017)

While a variety of harmful compounds, for example, cadmium, TPH and other chemicals can be discharged into the environment as a result of oil and gas companies’ operations. Many of these contaminants have been proven to be harmful to human health. Exposure to cadmium as well as hydrocarbon (benzene) is classified as very carcinogenic for humans by the International Agency for Research on Cancer. Additionally, groundwater pollution is probable to contain very harmful substances resulting from industrial production, such as lead, prior epidemiological research has indicated that main lead health outcomes include cancer as well as congenital malformation and have been found to be statistically associated with exposure to groundwater pollutions (Morufu and Clinton, 2017; Raimi and Sabinus, 2017b; Premoboere and Raimi, 2018; Olalekan *et al.*, 2018b; Olalekan *et al.*, 2020a; Deinkoro *et al.*, 2021; Morufu *et al.*, 2021a). Arsenic, chromium, fluoride, barium, boron, selenium as well as uranium are all common geogenic chemicals that have health implications in drinking water (WHO, 2011; Olalekan *et al.*, 2018b; Olalekan *et al.*, 2020a), nonetheless arsenic (As) as well as fluoride (F) have received the maximum attention owing to their toxicity as well as widespread distribution (Smedley and Kinniburgh, 2002; Deinkuro *et al.*, 2021). The geochemical features of the aquifer material, especially significant concentrations of the pollutant within the rock matrix may cause geogenic contamination of groundwater. This dissolves as a result of water-rock contact and/ or environmental conditions, making the pollutant more mobile. The aquifer type (whether unconfined or confined) as well as the depth of water table are two factors that influence naturally occurring pollutants. Unconfined aquifers remain more prone to pollution than confined aquifers, according to Berardinucci and Ronneseth (2002), since groundwater is younger as well as moves through the system more quickly. Surface derived contaminants remain more sensitive in aquifers with shallow water tables. Over 200 million individuals globally, according to UNESCO estimates, rely on drinking water that contains trace elements concentrations in amounts that exceed the current WHO (2017) recommendation (equal to the population of Nigeria, thus detrimental to life expectancy than COVID-19 in Nigeria). At the local, regional and national levels, access to clean drinking-water is a critical health as well as development concern (WHO, 2011; Olalekan *et al.*, 2019a; Raimi *et al.*, 2019a; Gift and Olalekan, 2020; Gift *et al.*, 2020; Olalekan *et al.*, 2020a). Controlling drinking-water quality necessitates the growth of management strategies that involve a drinking water assessment system toward identifying potential dangers. As groundwater becomes a significant supply of freshwater for residential use in the Niger Delta along with most Nigerian cities, it is critical to examine its quality assessment, particularly in terms of geogenic pollutants. This is because people’s reliance on groundwater from shallow aquifers puts a large number of individuals at contamination risk to geogenic sources. Trace elements remain amongst the few chemicals that have remained demonstrated to have extensive health problems in humans as a result of excessive amounts of exposure by drinking-water (WHO, 2011; Raimi and Sabinus, 2017b; Olalekan *et al.*, 2018b; Raimi, 2019; Olalekan *et al.*, 2020a), whereas iron is important due to its acceptability effects (Morufu and Clinton, 2017; Olalekan *et al.*, 2018b; Olalekan *et al.*, 2020a). As a consequence, towards addressing these concerns, the goal of this research paper is to compare differences in water quality parameters in the study region (determine the level of pollutions in the different sites) in the vicinity of “Gas Flaring Area of Ebocha-Obrikom of Rivers State, Nigeria”. Thus, this research will provide valuable information and add to our understanding on the physico-chemical examination of drinking water associated with the contamination of the ground waters by petroleum products. Hence, the ground water quality in the study area will be highlighted in this study: it will also bring to the awareness of the local people the type of water that is good for them as drinking water according to recommended standards. Also, it will provide a structural framework for effective and accurate management of groundwater and provide an available reference source and base line data for researchers involved in water resources assessment; thereby, contributing to the existing literature. The study will help government in policy intervention and supervision for better health service delivery. Furthermore, it will help in integrating the health needs of the populace into the state health scheme, in recognition of the fact that health is required for national development.

## 2. United Nations Sustainable Development Goals (UN SDGs) and Groundwater Pollution

Pollution from groundwater has been globally documented as a foremost threat to public/environmental health, as well as it obstructs the United Nations Sustainable Development Goals (SDGs) accomplishment, together with those associated to poverty eradication (SDG 1), zero hunger (SDG 2) as well as good health and wellbeing (SDG 3), ensure access to clean water and energy, address needs for quality education and tackle climate change, and protect life on land and in the oceans. They address humanity’s challenges, and are a universal call to protect the planet and improve lives. To support this process, drinking safe water is a basic necessity for human development, health as well as well-being, and hence, an internationally recognized human right and has long been a goal of national and international policy (WHO, 2001; Olalekan *et al.*, 2018a; Raimi *et al.*, 2019a; Olalekan *et al.*, 2019a; Gift and Olalekan, 2020; Gift *et al.*, 2020; Olalekan *et al.*, 2020a). Pollution from groundwater disproportionately affects the most disadvantaged, particularly children as well as women (SDG 5) (Raimi *et al.*, 2019b; Raimi *et al.*, 2019c; Morufu *et al.*, 2021a). Contaminants leaching into groundwater as well as runoff threatened the availability of safe drinking water (SDG 6) (see table 1 below). The Sustainable Development Goals (SDGs) of the United Nations comprise ambitious worldwide targets for drinking water, sanitation as well as hygiene. The SDG indicator for target 6.1, usage of safely managed drinking water services (SMDW), attempts to rectify the shortcomings of prior monitoring efforts. Improved drinking water sources (protected groundwater sources, piped water, rainwater collection as well as packaged or delivered water) that remain available on premises, accessible when required as well as free of contamination are described as SMDW services. Thus, the need to increase drinking water quality monitoring in Nigeria’s Niger Delta Region is widely recognized (Morufu and Clinton, 2017; Raimi and Sabinus, 2017b; Olalekan *et al.*, 2018b; Olalekan *et al.*, 2019a; Olalekan *et al.*, 2020a; Olalekan *et al.*, 2021). Oil and Gas industry domicile in the area may be positioned to include water quality monitoring in their activities; as it is remarkable that water safety activities do not compromise. While, the UN Sustainable Development Goals (SDGs) remained introduced in 2015 as a broadly agreed, wide-ranging plan of action aimed at environmental sustainability, social inclusion, as well as economic development. Because the extractive industry plays such a significant role in the universal economy, the SDGs are primarily targeted at governments. The 17 Sustainable Development Goals (SDGs), defined by the United Nations (UN) in its 2030 Agenda for Sustainable Development offer a framework/roadmap aimed at creating a better future for people as well as the planet come 2030. The SDGs were adopted by all United Nations Member States in the year 2015 and remain a call to action for all countries – rich, middle-income and poor, to increase prosperity while safeguarding the milieu. They understand that water is a vital resource for agriculture, human development, industry and that it can contribute to economic progress and meet a variety of societal demands (Suleiman *et al.*, 2019; Raimi *et al.*, 2019a; Raimi *et al.*, 2019d; Olalekan *et al.*, 2021; Morufu *et al.*, 2021a). The United Nations (UN) considers water access as well as sanitation to remain a human rights problem, with everyone entitles to adequate, acceptable, safe, physically-accessible and inexpensive water for personal as well as household purposes (Olalekan *et al.*, 2018a; Raimi *et al.*, 2019a; Olalekan *et al.*, 2019a; Raimi *et al.*, 2019d; Gift *et al.*, 2020; Gift and Olalekan, 2020). Demand for freshwater resources is anticipated to intensify as the world’s population grows, urbanization accelerates together with agricultural and economic development, (Isah *et al.*, 2020a; Olalekan *et al.*, 2020b; Isah *et al.*, 2020b; Hussain *et al.*, 2021a; Morufu, 2021; Hussain *et al.*, 2021b; Morufu *et al.*, 2021b). Changing climate, land use, as well as water availability, reliability and quality are just a few of the concerns that could have an impact on the activities of the oil as well as gas industry (Raimi *et al.*, 2018a; Adindu and Raimi, 2018; Olalekan *et al.*, 2019a; Ihuoma and Raimi, 2019; Adindu *et al.*, 2019; Henry *et al.*, 2019b; Ebuete *et al.*, 2019; Raimi *et al.*, 2018b; Omidiji and Raimi, 2019; Raimi *et al.*, 2019e; Adedoyin *et al.*, 2020; Raimi *et al.*, 2020a; Raimi *et al.*, 2020c; Olalekan *et al.*, 2020c; Olalekan *et al.*, 2020d; Morufu *et al.*, 2021c; Raimi *et al.*, 2021b). For example, industry operators might consider locating operations in areas where water supply and quality are presently or may become a problem in the future, or in areas prone to extreme weather as well as flooding (Odubo and Raimi, 2019). Water scarcity can have a remarkable consequence for local populations as well as stakeholders (Olalekan *et al.*, 2019b). Industrial users, such as the oil and gas industry, may face physical, regulatory as well as reputational issues as a result. As a result, the International Petroleum Industry Environmental Conservation Association (IPIECA) and others collaborated an Atlas that depicts the oil and gas industry contribution to the SDGs, with all SDGs potentially relevant. Figure 2 depicts the guidance modules, which provide particular information that can be used to illustrate SDGs contribution.

**Figure 2:**
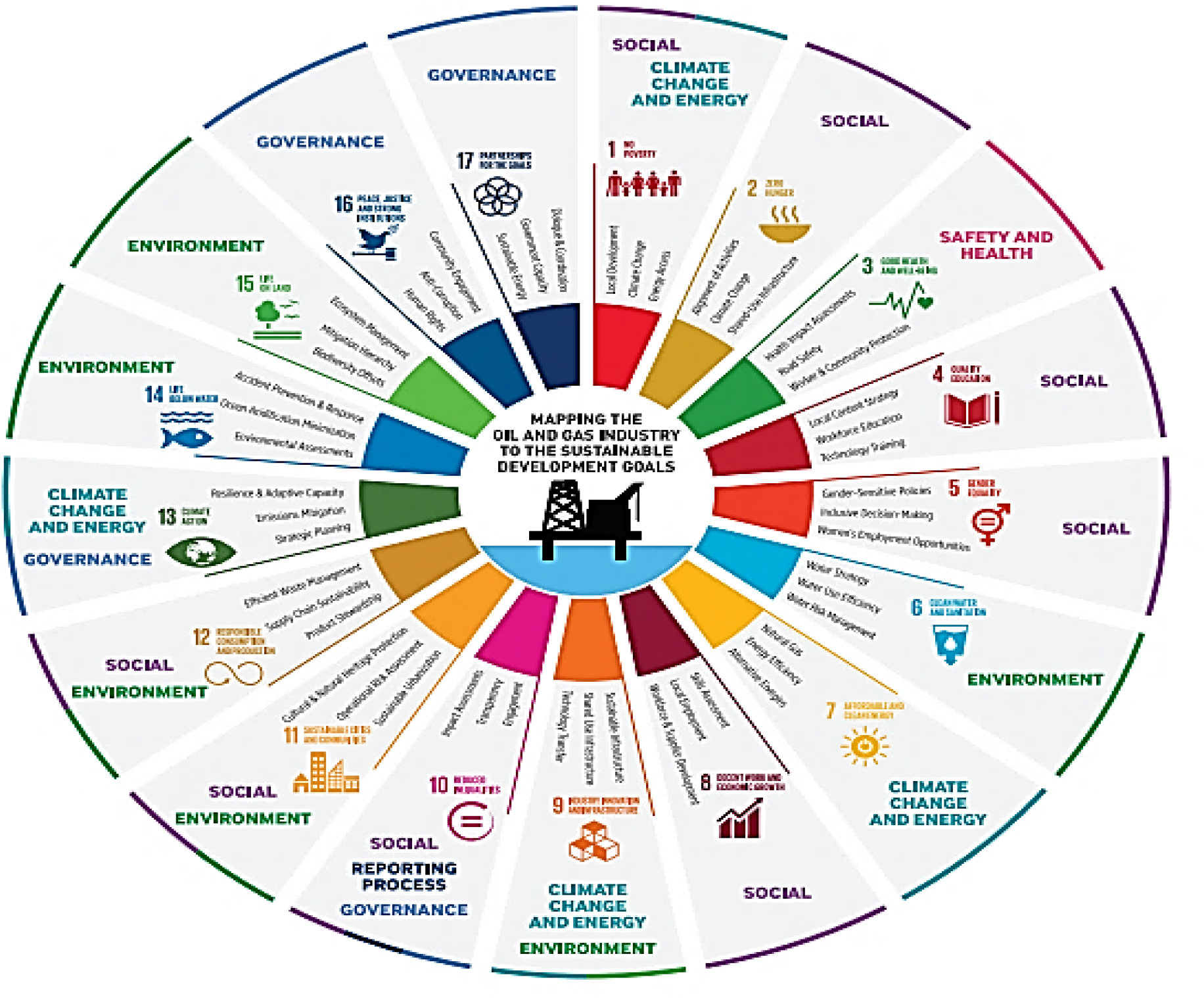
Mapping the modules to the SDGs. Adapted from Global Reporting Initiative and United Nation (2015)

**Table 1.**
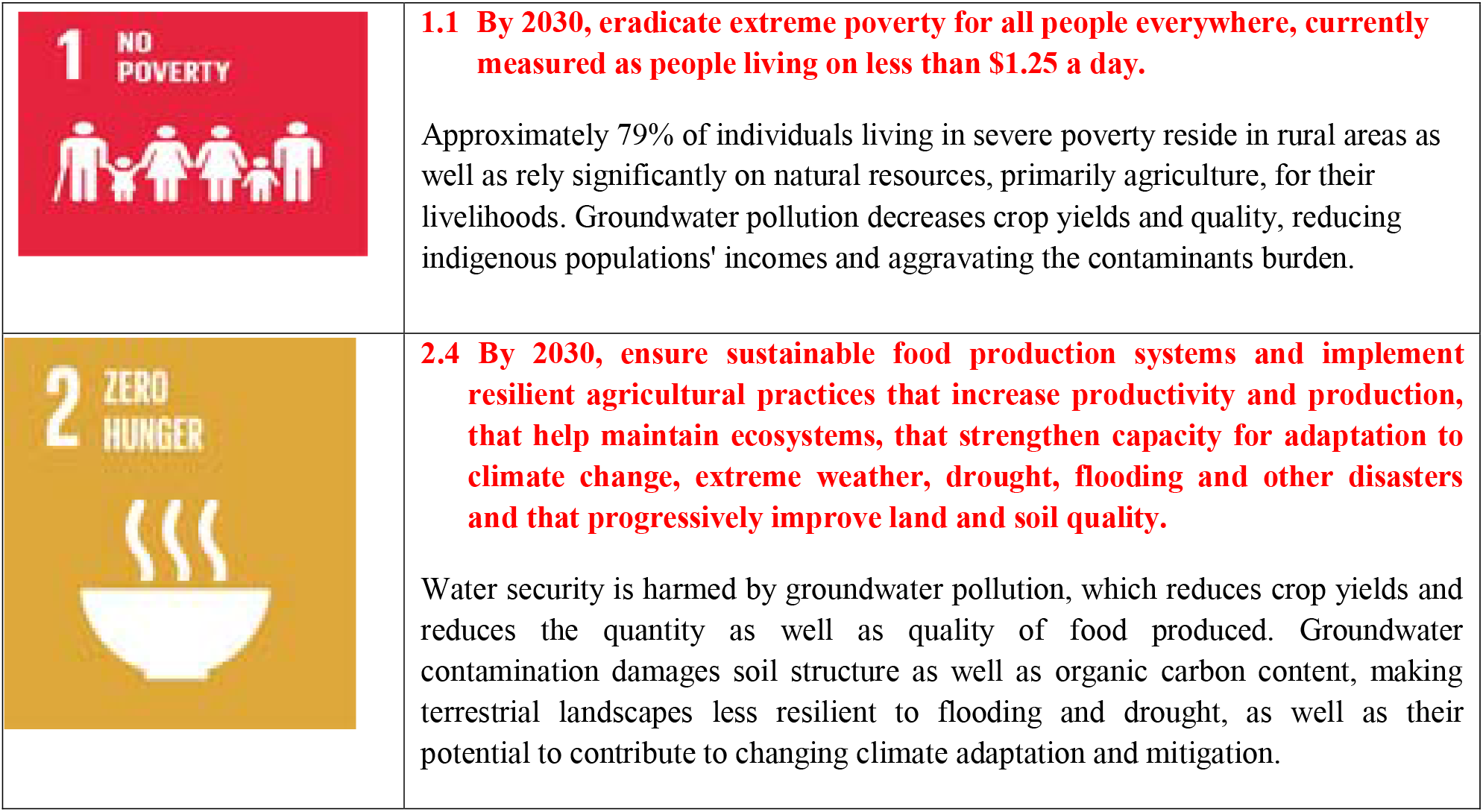
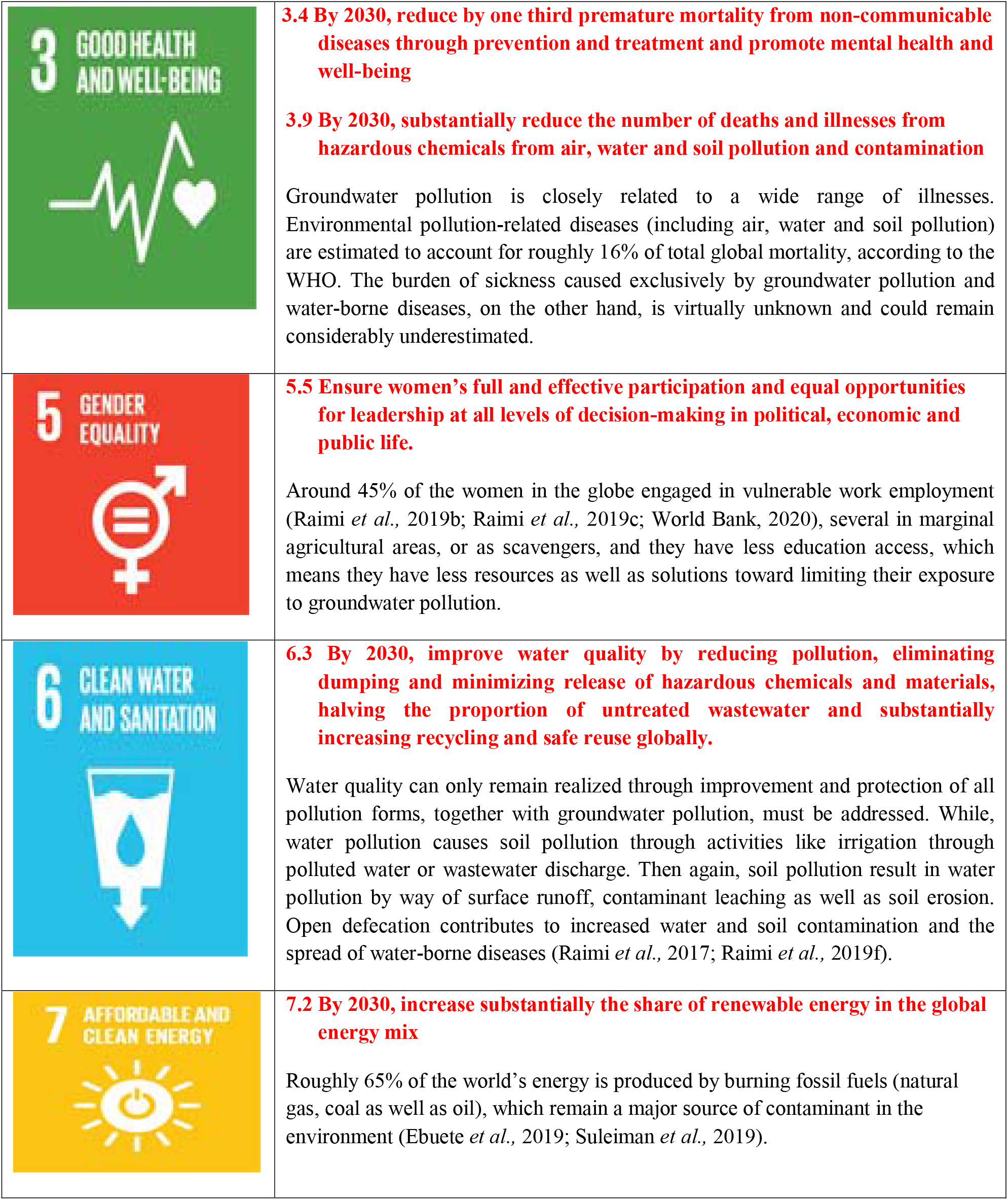
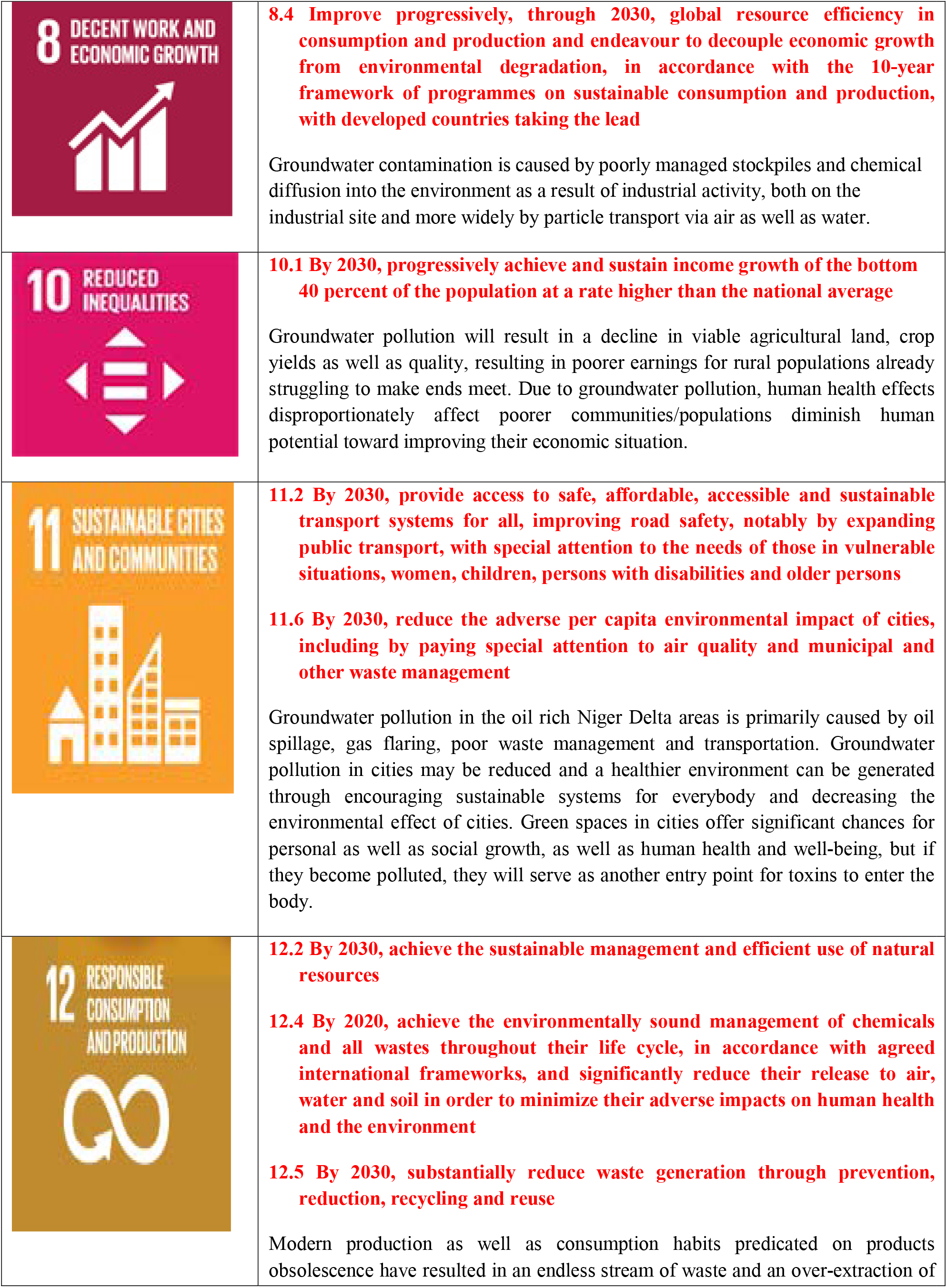
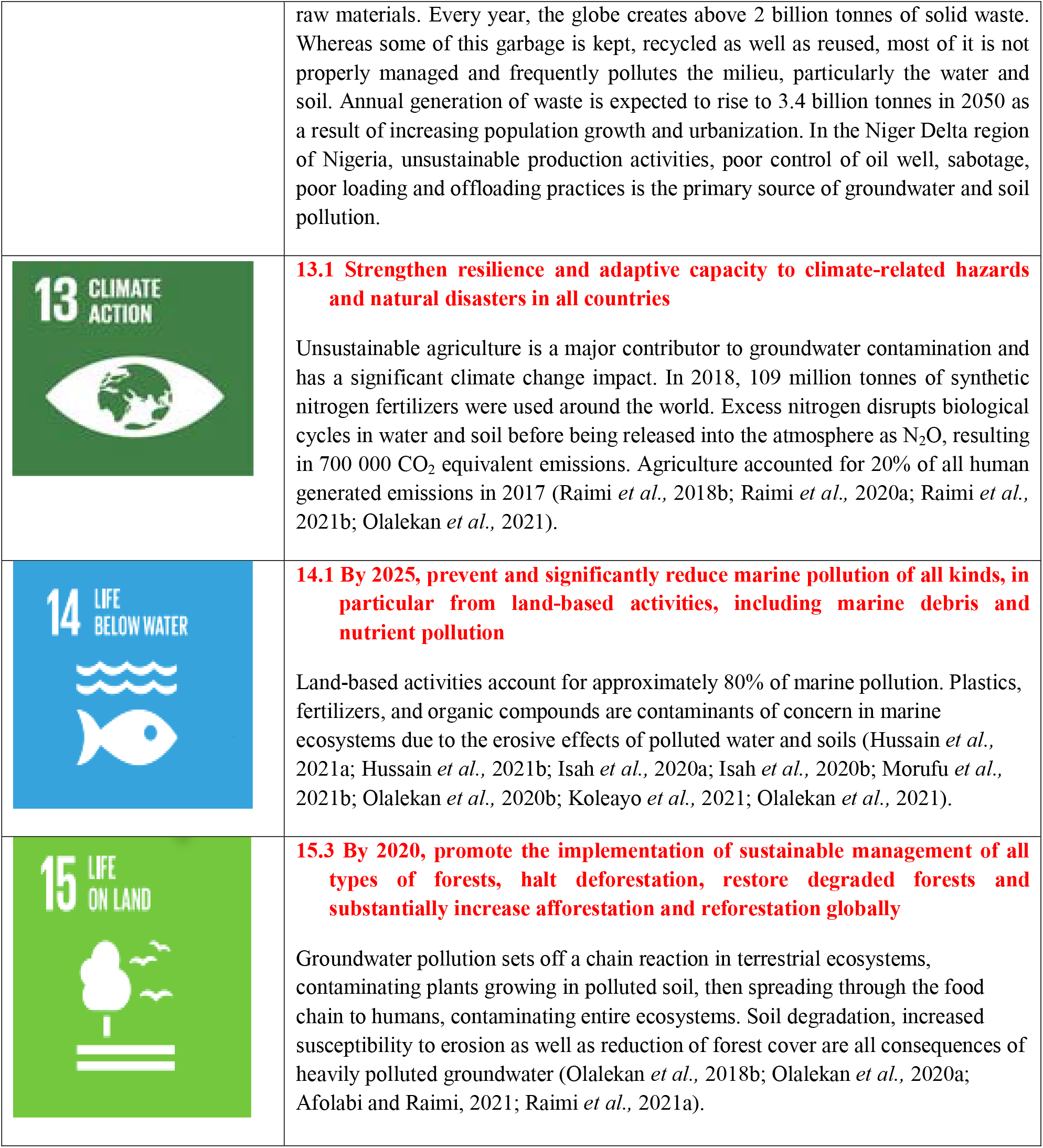
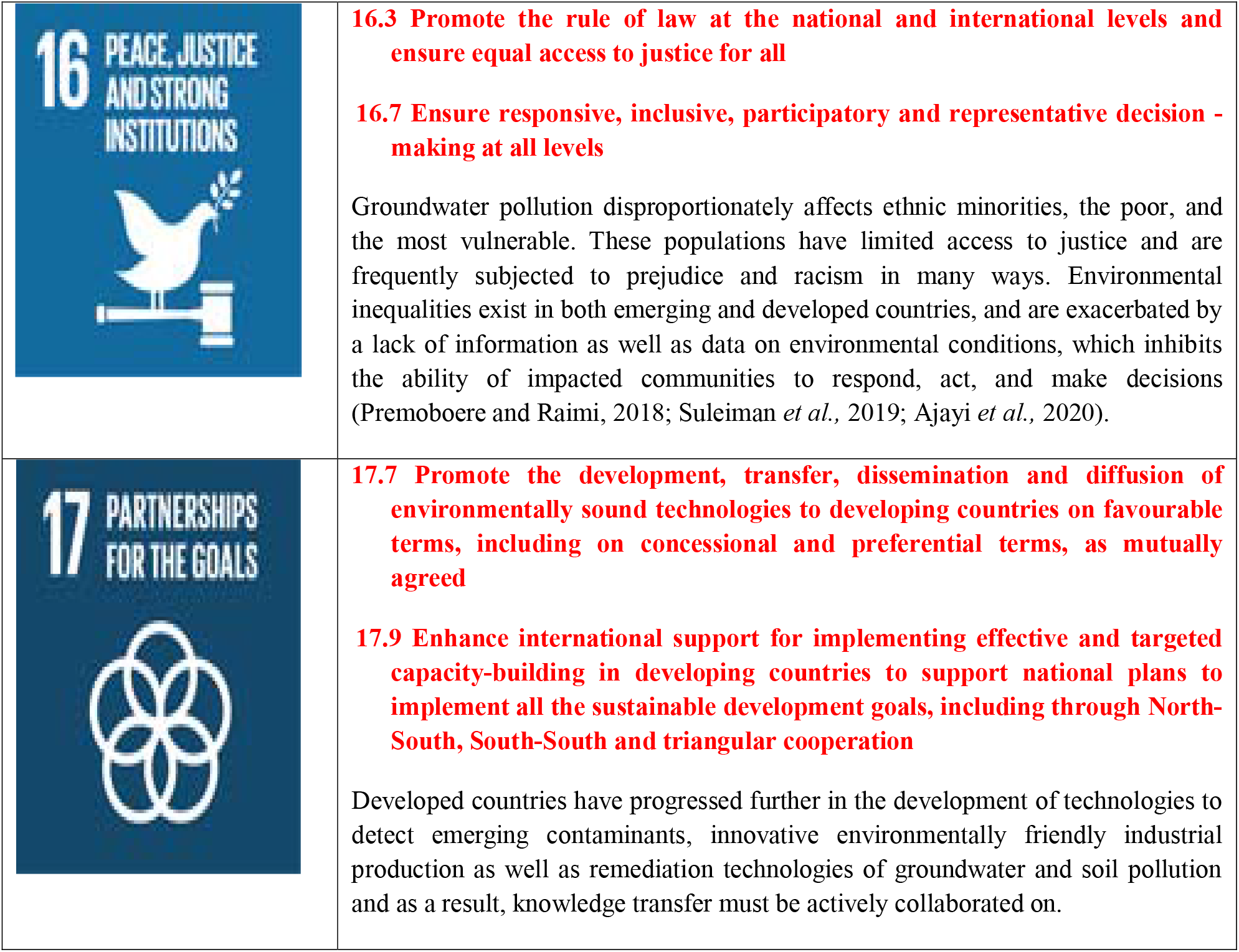
Links between Sustainable Development Goals and Groundwater Pollution.

Hence, the complexity of the SDGs means that many of the goals need to be addressed through social, political and financial changes. Science and technology are vital to this process, according to the 2016 Global Sustainable Development Report (https://sustainabledevelopment.un.org/content/documents/10789Chapter3_GSDR2016.pdf). Advances in nanotechnology could help improve battery storage or water filtration; developments in Earth sciences could inform natural resource policy; and breakthroughs in medicine could impact healthcare. Addressing the SDGs therefore requires interdisciplinary research: collaboration among natural and social scientists, policymakers, and industry leaders.

## 3. Conceptual Framework

From extraction through end-product supply, the oil and gas sector comprise a wide range of activities. The value chain is the term used to describe this spectrum. Figure 3 depicts the kind of activities that a fully “integrated” oil and gas corporation with broad upstream and downstream operations might engage in. Thus, an oil and gas activities from companies at a particular area can span decades, for example, from early offshore exploration to platform decommissioning. Hence, there is need for companies to be encouraged to evaluate the effects of their actions across the various value chain.

**Figure 3:**
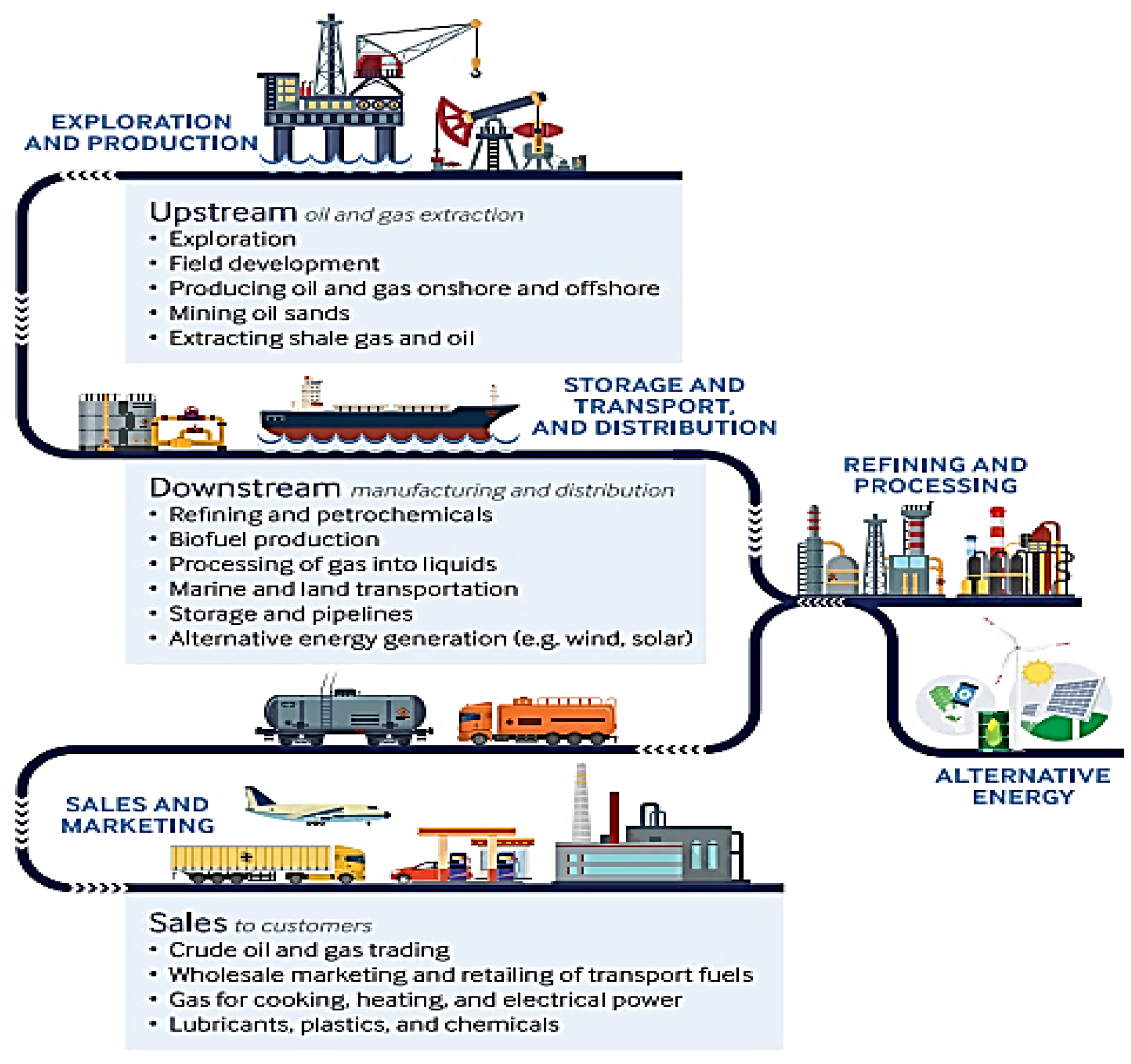
Oil and gas value chain. Adapted from IPIECA, (2017)

While, the Niger Delta region of Nigeria produce majority of the nation’s oil and gas, producing 85 million estimated barrels of oil equivalent daily (Premoboere and Raimi, 2018; Deinkuro *et al.*, 2021; Morufu *et al.*, 2021a). Despite the fact, oil companies operating in the Niger Delta have an impact on how millions of individuals gain (or suffer) from the country’ hydrocarbon reserves. These corporations oversee “multi-billion-dollar portfolios of public assets”, carry out complicated projects across their territories as well as at sea, employ tens or hundreds of thousands of individuals, as well as provide a variety of public services ranging from energy generation to infrastructure building. Regardless of their relevance, flaring of natural gas and oil spills may have severe as well as multiple environmental, health, social as well as economic effects on a company’s reputation and indigenous communities (Premoboere and Raimi, 2018; Suleiman *et al.*, 2019; Olalekan *et al.*, 2020a; Deinkuro *et al.*, 2021; Morufu *et al.*, 2021a). Oil spills, which are likely to occur as a result of operational incidents, inadequate maintenance, or corrosion of equipment, can have serious as well as widespread environmental, social, health and economic effects in the worst-case scenario (Premoboere and Raimi, 2018; Ebuete *et al.*, 2019; Suleiman *et al.*, 2019; Olalekan *et al.*, 2020a; Deinkuro *et al.*, 2021; Morufu *et al.*, 2021a). Spills can also have serious long-term ramifications for a company’s reputation. The health risks and impacts from oil gas industry resulting from groundwater pollution remain linked to some contaminants (or hazardous substances) created by physical-chemical interactions. Contaminants can travel through the soil, air as well as water in general (Raimi *et al.*, 2019a; Raimi *et al.*, 2021a; Olalekan *et al.*, 2021). They can likewise settle on or be digested through animals or plants and they can end up in the food chain, air as well as water (Ajayi *et al.*, 2020; Olalekan *et al.*, 2020a; Afolabi and Raimi, 2021). The various ways in which an individual can come into contact by means of contaminants remain referred to as exposure pathways. There are three primary routes of exposure, namely: ingestion, inhalation as well as skin contact. Inhalation is the act of breathing or inhaling into the lungs. Ingestion is the act of ingesting something via mouth. When anything comes into direct contact with the skin, this is referred to as skin contact. After skin contact, ingestion can be a secondary exposure pathway. Acute or chronic exposures can occur. An acute exposure is defined as a single brief exposure to a harmful material (pollutant). Health symptoms may emerge quickly following exposure; for example, a gas release is hazardous because to the risk of fire and explosion, whereas an oil release might have very different and potentially catastrophic implications for the environment and individuals. Thus, leading to a cause, life loss, or a serious fire. Prolonged exposure happens over a significantly longer length of time, frequently with smaller amounts of exposure in repeated ways. People living near Love Canal (Goldman *et al.*, 1985), a leaking hazardous waste dump, for instance, did not recognize the health repercussions of prolonged exposure for some years. Chronic health impacts are often long-term illnesses or injuries like liver failure, cancer, stunted growth as well as development. Bioaccumulation is one of the mechanisms by which persistent exposure to even trace levels of harmful substances can cause harm. Some compounds are absorbed rather than expelled and remain in the human body. They build up and cause harm over time. As a result, detrimental health impacts are determined by the determinants of exposure. The following factors influence whether or not unfavorable health impacts may occur as a result of an exposure:

○ The pollutant nature;
○ The dosage or amount (the amount or pollutant level indigenous population or an individual was exposed to);
○ The extent (how long did exposure last);
○ The regularity (how frequent did the individual was exposed).

As a result, any attempt to link groundwater pollution to health risks as well as impacts will almost probably include an assessment of the following criteria (Raimi and Sabinus, 2017b; Olalekan *et al.*, 2018b; Olalekan *et al.*, 2020a; Morufu *et al.*, 2021a):

○ The mass rate of pollution emission from both waterborne as well as airborne sources.
○ The contamination level on a regional scale, as well as the persistence and the pollutants transformation including the consequent products of their transformation.
○ Pollutants concentrations as well as gradients that adversely influence water, air as well as land resources.
○ The number of individuals, particularly sensitive groups who may be impacted by the pollutant’s emission from the gas flaring site.
○ Total amount of time spans that pollutant is released: Contaminant(s) as well as concentrations - Throughout their lifetimes, humans remain exposed to a variety of contaminants. The contaminants mixtures to which we remain exposed change throughout our lives.
○ Exposure duration.
○ The synergistic as well as antagonistic or additive effects of additional pollutant releases unfavorable health conditions that may make an exposed population more pollutants sensitive through the gas flaring site.
○ Gas flaring site characteristics like the depth and degree of pollution.
○ The total amount disposed of and the areal coverage define the gas flaring site size.

The entire procedure of health risks assessment and the groundwater impacts is quite challenging and necessitates a great deal of expertise, time as well as financial resources to complete. Its successful implementation necessitates dealing with the lack of exact data on the dosage response association for some of the concern chemicals, as well as making a variety of educated assumptions as well as interpretations in order to gain an improved understanding of what is more or less essential. For the study implementation, environmental samples (water and soil) need to be examined toward determining the content as well as several pollutants concentrations (polychlorinated biphenyls, heavy metals and pesticides) known to be harmful to human health. Groundwater samples collected from the gas flaring site were compared to samples collected from another location, a peri-urban residential area approximately 30km away from Ebocha-Obrikom oil and gas field. Below, is the flow chart (figure 4) of the study, highlighting the relationship amongst the environmental contaminants from the groundwater pollution as well as environmental/public health impacts on the indigenous and surrounding communities. This flow chart, which shows the conceptual relationship between health and groundwater pollution, is typical of any similar investigation. One of the most critical variables that determine the public/environmental health dangers of groundwater is the waste streams disposed of in it.

**Figure 4:**
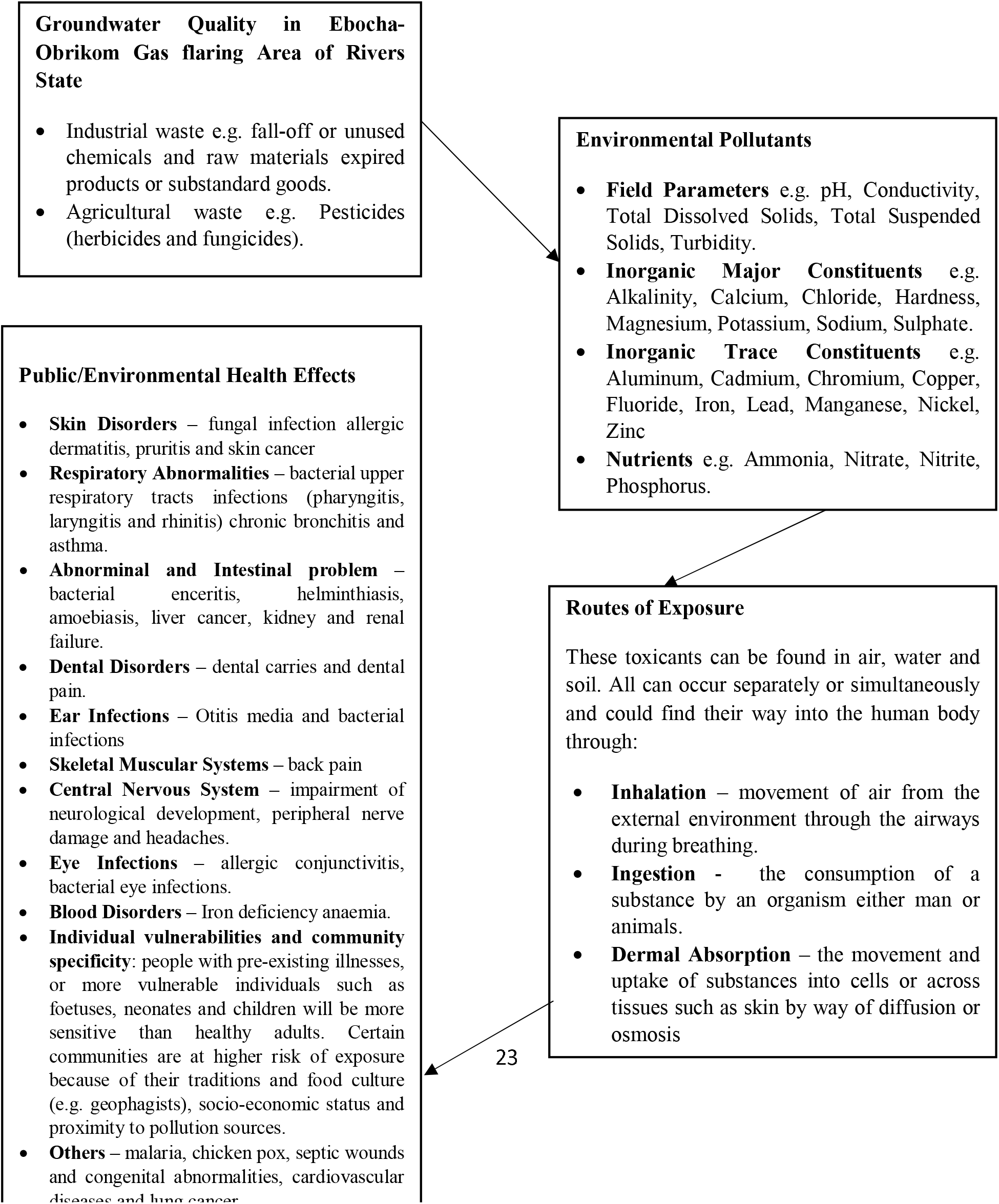
The flow chart of the Groundwater Quality in Ebocha-Obrikom Gas flaring Area of Rivers State showing the conceptual framework between exposure to groundwater pollution and health.

## 4. Material and Methods

### 4.1 The Study Area

The area is placed between latitude 5^0^ 20N - 5^0^ 27N and longitude 6^0^ 40E - 6^0^ 46E is situated in Ebocha-Obrikom (Figure 5). It encompasses Obrikom, Obie, Obor, Ebocha as well as Agip New Base towns all located in Ogba/ Egbema/Ndoni Area of Rivers State (Figure 5). The study research area lies to the North by River Nkissa, by the West, River Orashi, by the East, River Sombrero and by the South Omoku town (Morufu *et al.*, 2021a).

**Figure 5:**
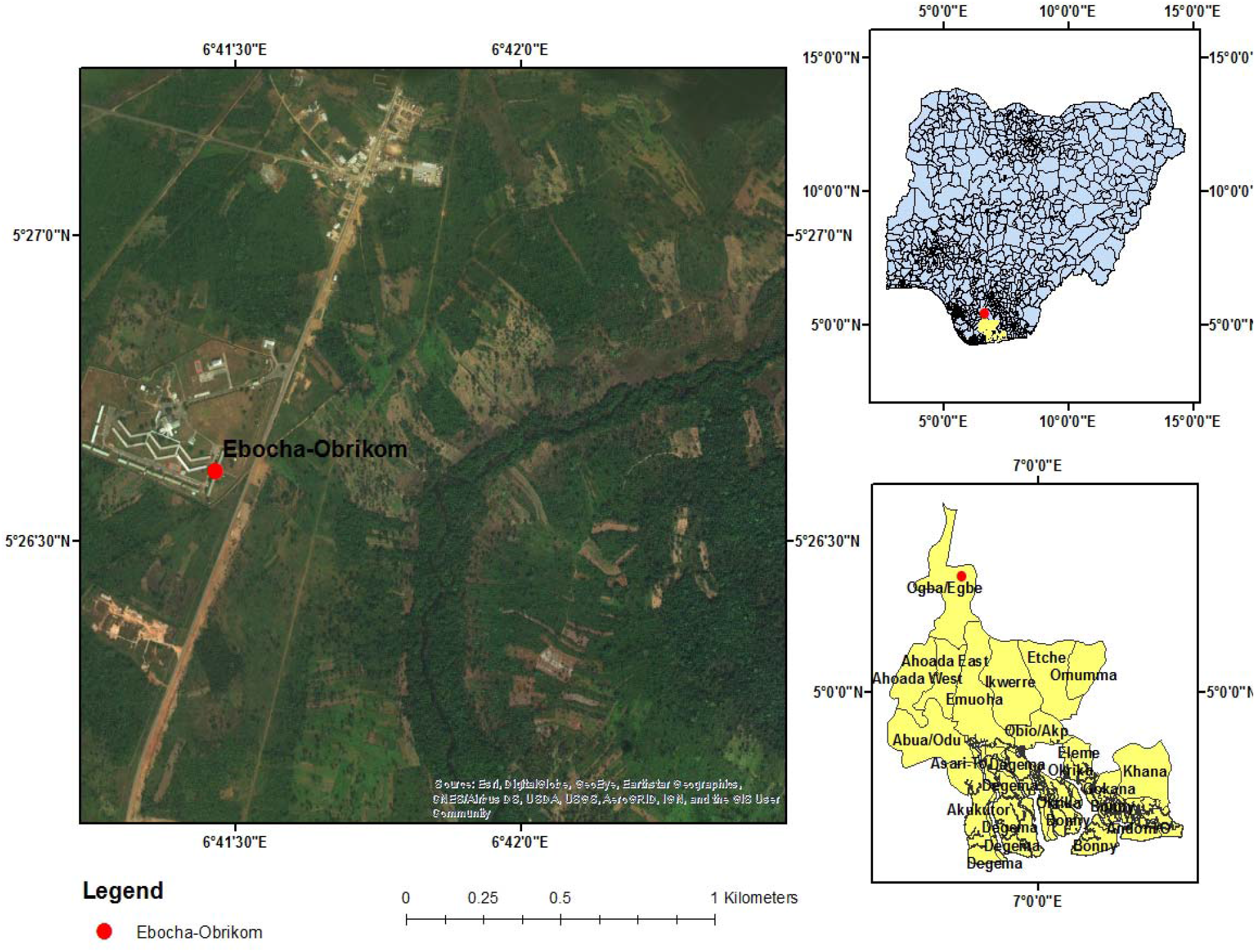
Map Showing the Study Area with Nigeria and River State insert. **Sources:** Adapted from Olalekan *et al.*, 2018b [https://doi.org/10.4236/ojogas.2018.33017]

### 4.2 Climate

The climate of the study area is the equatorial type. Heavy rainfall of about 2500 mm/annum are experienced in the region. During the year, the rainfall takes around eight (8) months (March to October), and even months deemed dry remain often not devoid of occasional precipitation (Morufu and Clinton, 2017; Raimi and Sabinus, 2017b; Olalekan *et al.*, 2018b; Morufu *et al.*, 2021a). Temperature is usually high and the mean monthly temperature is about 25^0^C (Morufu and Clinton, 2017; Raimi and Sabinus, 2017b; Olalekan *et al.*, 2018b; Morufu *et al.*, 2021a).

### 4.3 Vegetation

The vegetation is dominated by fresh water swamps, spotted areas are characterized by stratified high forest: The vegetation comprises a multitude of evergreen trees that yield tropical hard wood, e.g., mahogany. Smaller palm trees are present, ascending plants such as lianas or rattan palms may be hundreds of meters long and also epiphytes and parasites that grow on other plants. A vast variety of orchids, creepers and ferns thrive below the trees (Morufu and Clinton, 2017; Raimi and Sabinus, 2017b; Olalekan *et al.*, 2018b; Morufu *et al.*, 2021a). The traditional rainforest vegetation has been replaced in most instances by settlements, farms, fallow and secondary forests, construction of civil structures, oil and natural gas exploration/exploitation. The only areas where primary forests may be found are raffia palm dominated riparian forests along the Sombreiro River, across the Orashi River and sacred groves which is relic of traditional African religion (Morufu and Clinton, 2017; Raimi and Sabinus, 2017b; Olalekan *et al.*, 2018b; Morufu *et al.*, 2021a).

### 4.4 Topography and Drainage

The area is actually drained by the Sombreiro on the Eastern flanks and Orashi on the Western flank creating an almost uninterrupted inter-basinal area. The region has a topographic structure virtually flat, as well as is covered by superficial soil, which is made up of silty clays combined with silty sand. There is a clear lack of imposing hills that rise above the whole land surface at a height of about 25 metres above sea level (Morufu *et al.*, 2021a).

The Orashi River is a prominent feature of the natural drainage system. This river, although an independent river system, which accounts for the drainage of the entire zone, links up with the Niger Delta system during the wet season (flood stage). Due to the more prominent relief of this area, drainage is more efficient and much fewer rivers and creeks drain the area. Backswamp depressions exist which entrap floodwaters and so form perennial lakes in the area. The rivers are prone to flooding which increases the level of water in the water table. The rivers found in the study area are also subjected to tidal flow (Morufu and Clinton, 2017; Raimi and Sabinus, 2017b; Olalekan *et al.*, 2018b; Morufu *et al.*, 2021a).

### 4.5 Soil

The soil occurring on this area is part of the recent alluvium of the Niger Delta and shows age differences in a better development of Argillic horizon (clay illuviation). They are dominantly sandy loam on top soil changing to sandy clay loam and then clay in the subsoils. The soil reaction is acid (pH 4.3-5.0) (Ogoni, 2010). The high acidity of the soils is due to the high Aluminium (Al) content of the soils (Alagoa, 2005; Morufu and Clinton, 2017; Raimi and Sabinus, 2017b; Olalekan *et al.*, 2018b). The soil organic matter content is low as well as could be due to continuous cultivation of the land. The organic matter decreases down the soil profile. Available phosphorus (P) is moderate especially in middle horizons. The soil fertility in term of percent base saturation is high. The soils are mainly Alfisols (Luvisols and Natrosol), Ultisols (Regosols) and Oxisols (orthic Ferralsols). Other soils identified are inceptisols (Gleyic cambisols) and entisols (Albic Arenosols). The soils are Ultisols (Eutric-Paleudalfs) (Alagoa and Derefaka, 2002; Morufu and Clinton, 2017; Raimi and Sabinus, 2017b; Olalekan *et al.*, 2018b).

### 4.6 Land Use

The Ebocha-Obrikom area has various patterns of land use. The only perceptible activity of economic importance in the environment is the crude petroleum exploration as well as exploitation. The land use pattern of the area is basically characterized by a number of agricultural activities which include farming, as well as fishing activities which are among the major activities in the area. Some of the crops cultivated include cocoyam, okra, bitter-leaf, cassava, yam etc. The oil palm planting harvesting and processing are also important (Alagoa, 2005; Morufu and Clinton, 2017; Raimi and Sabinus, 2017b; Olalekan *et al.*, 2018b; Morufu *et al.*, 2021a).

### 4.7 Sample Collection

The sampling strategy employed for the current research investigation were similar to that utilized by Morufu and Clinton (2017); Raimi and Sabinus (2017b); Olalekan *et al*. (2018b) in which sampling was targeted in some vulnerable quarters at a densely populated location. These quarters are places predisposed to all kinds of contamination not only because of their geographical situation but also because of the presence of crude petroleum exploration and exploitation. From the sample location (see table 2 below), extracted water samples from groundwater sources utilized mostly for drinking as well as domestic activities. Sample collections were limited to only groundwater from dug wells or shallow pumping wells built for household uses exclusively. The depth of the wells varies between 10 to 28m, which is a phreatic aquifer. The sampling locations sites were documented using portable GPS devices. In the vicinity of the depot, ground water sources were selected randomly but at various distances from each other for the purpose of this investigation. Furthermore, the samples were manually collected from nine (9) strategic locations in the study area for ground water (boreholes and wells) into previously washed clean plastic sampling bottles after about 20 min of continuous water flow to ensure adequate aquifer quality that can be appropriately represented.

**Table 2.** Geographical coordinates of the nine (9) sampling sites (samples).

| **S/N** | **Locations** | **Altitude (m)** | **Latitude** | **Longitude** |
| --- | --- | --- | --- | --- |
| Site – 1 | (Borehole) (Opposite Ijeoma Quarters. 750m Away from Agip Gas Flaring Center Ebocha) | 10 | Lat N05<sup>0</sup> 27' 068" | Long E006<sup>0</sup> 41' 480" |
| Site – 2 | (Borehole) (200m Opposite Agip Gas Flaring Centre Ebocha and 50m from Agip Waste Pit) | - | Lat N05<sup>0</sup> 27' 28.7" | Long E006<sup>0</sup> 41' 58.1" |
| Site – 3 | (Well) (The Apple Hotel 500m from Waste Pit and 150m Away from Mgbede Field Oil Well 7 Ebocha) | 16 | Lat N05<sup>0</sup> 27' 37.5" | Long E006<sup>0</sup> 42' 05.3" |
| Site – 4 | (Well) (1000m Away from the Agip Flare Stack Ebocha) | 22 | Lat N05<sup>0</sup> 26' 51.5" | Long E006<sup>0</sup> 41' 38.8" |
| Site – 5 | (Borehole) (Abacha Road Obrikom, 800m Away from Agip Gas Plant) | - | Lat N05<sup>0</sup> 23' 48.6" | Long E006<sup>0</sup> 40' 36.8" |
| Site – 6 | (Borehole) (Eagle Base Obor. 2,500m Away from Agip Gas Plant) | 28 | Lat N05<sup>0</sup> 23' 00.9" | Long E006<sup>0</sup> 41' 07.4" |
| Sites – 7 | (Well) (Obor Road Obie. 2000m Away from Agip Gas Plant) | 24 | Lat N05<sup>0</sup> 23' 22.5" | Long E006<sup>0</sup> 40' 49.1" |
| Sites – 8 | (Borehole) (Green River Plant Propagation Centre Naoc 3000m Away from Agip Gas Plant) | 17 | Lat N05<sup>0</sup> 24' 18.9" | Long E006<sup>0</sup> 40' 55.0" |
| Sites – 9 | (Control) (35,000m from Ebocha) | - | Lat N5<sup>0</sup> 4' 58.1412" | Long E6<sup>0</sup> 39' 30.4806" |

All of the samples was obtained during the daytime, from 9.00am to 4.00pm. Due of insecurity, flooding and COVID-19 lockdown. Night samples were not collected and sampling was performed between September, 2019 to August 2020.

### 4.8 Sampling, Preservation and Analysis

The standard methods outlined in APHA (2012); Morufu and Clinton (2017); Raimi and Sabinus (2017b); Olalekan *et al*. (2018b); Morufu *et al*. (2021a) have been followed by water sampling, conservation, transportation as well as analysis.

### 4.9 Ground Water Collection

For the analyses of physico-chemical parameters, ground water samples were collected using pre-rinsed 1litre plastic containers. Pre-rinsed ground water samples for heavy metal analyses were collected with nitric acid of 1litre containers as well as treated with 2ml nitric acid (assaying 100%, Trace Metal Grade, Fisher Scientific) prior to storage. These were done to stabilize the metals oxidation conditions. Groundwater samples were collected in two groups of 250ml glass-stoppered-reagent bottles per sampling location for Biological Oxygen Demand (BOD), and Dissolved Oxygen (DO) determinations. The BOD samples have been properly filled without air trapping as well as the bottles covered in black polythene bags. This was done to eliminate light, which is present in the samples and capable of producing DO by autotrophes (algae). The BOD samples were incubated for five days, which was added to 2ml of each sample. In order to retard additional biological activities, Winkler solutions I and II use different dropping pipettes to each sample. The bottles were thoroughly shaken to precipitate the floc, which lay at the bottom of the bottles. Further, Winkler solution I is a solution of manganese sulphate, while solution II is sodium or potassium iodide, sodium or potassium hydroxide, sodium azide (sodium nitride) and sodium hydroxide. The DO samples were collected in clear bottles and also tightly stoppered. With samples of dissolved oxygen preserved on the spot with Winkler I and II solutions similar to that of the BOD samples (APHA, 2012). All samples had been clearly identified and controlled at 4°C for easy identification. Determination was carried out on site to know the concentrations of unstable as well as sensitive water quality characteristics including total dissolved solids (TDS), electrical conductivity (EC), pH, alkalinity (Alka.), and temperature (Temp). Thus, the fundamental approaches for investigating the groundwater composition are described in figure 6 below.

**Figure 6:**
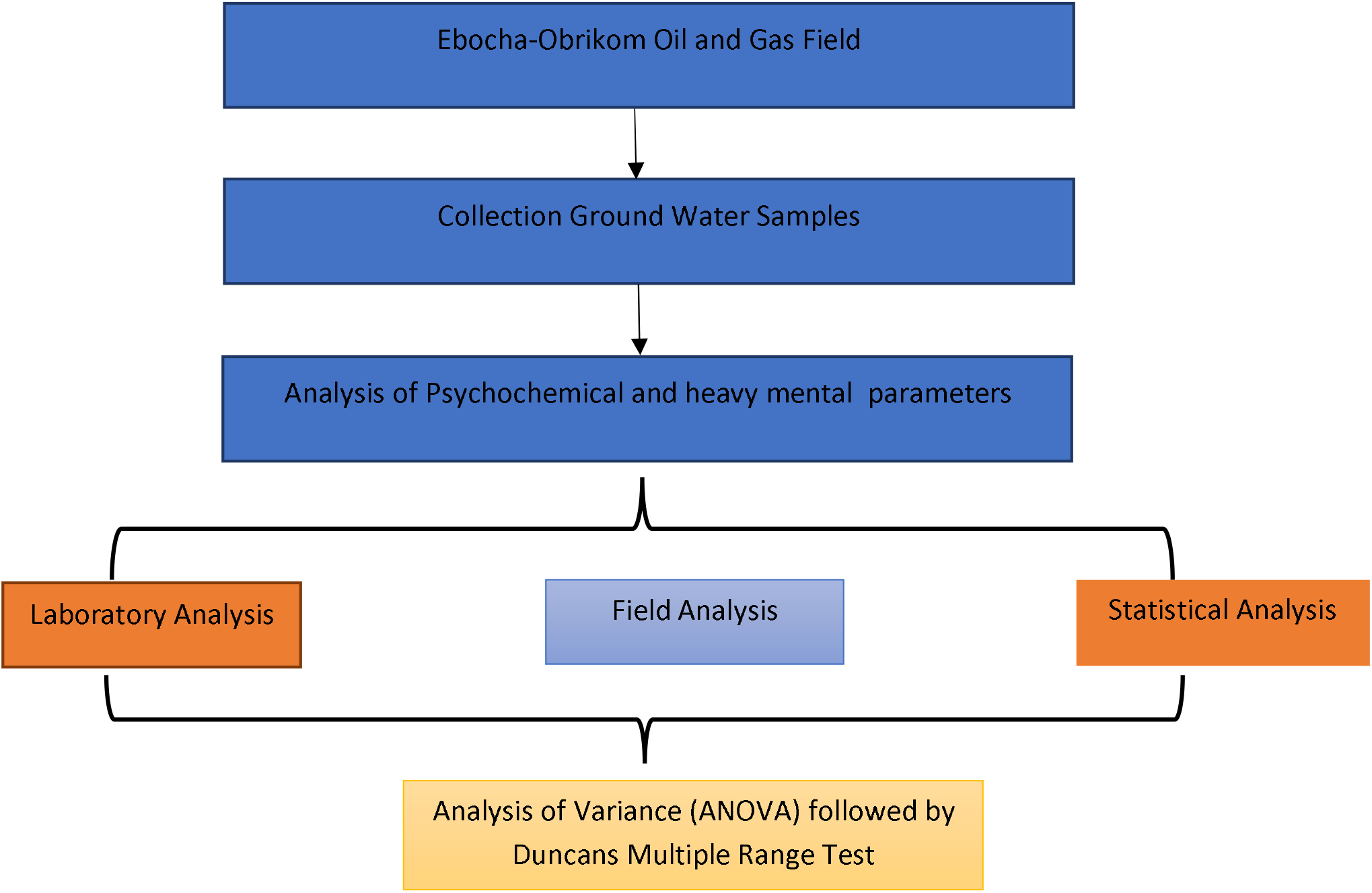
A schematic illustration of quantification methodology adopted for the current study.

### 4.10 Quality assurance and quality control (QA/QC)

Additionally, employing high purity analytical reagents as well as solvents, all analytical procedures were closely monitored with quality assurance as well as control techniques. Calibration standards were applied to the instruments. Procedure blanks, triplicate analysis as well as the analysis of certified reference materials (CRM) was performed through the analytical technique validation. For every organic pollutant from the groundwater samples, the limit of detection (LoD), repeatability, reproducibility, precision, as well as accuracy was established.

## 5. Results

### 5.1 Compare differences in water quality parameters in the study area (determine the level of pollutions in the different sites)

Results of site difference in water quality parameters in wet and dry seasons are presented in Tables 3 and 4. Result shows that during wet season, the mean values obtained for water quality parameters were significantly lower in site 9 compared with that obtained in other sites (p<0.05) with the exemptions of temperature, DO, BOD, COD, acidity, TH, TDS, K, Mg, Zn, Mn, Cd, Pb, Cu, Cr, NH_3_, NO_2_, NO_3_, Ni though slightly lower in most cases in site 9 were not significantly different (p>0.05) and both alkalinity and SO_4_ which were significantly higher in site 9 than site 1 (p<0.05) (Table 3). Result obtained during dry season reveals that there is no significant difference in pH, acidity, Pb and Ni between the nine sites (p>0.05) while other water quality parameters were significantly lower in site 9 than other sites excluding Cl and Mg which were both significantly higher in site 9 than site 8 (p<0.05) (Table 4).

**Table 3:** Comparison of the parameters in the different locations during raining season. Similar superscript means not significantly different (p>0.05) while different superscripts indicates significantly difference in means (p<0.05).

|  | Location |  |  |  |  |  |  |  |  |
| --- | --- | --- | --- | --- | --- | --- | --- | --- | --- |
| Parameters | 1 | 2 | 3 | 4 | 5 | 6 | 7 | 8 | 9 |
| Temperature (<sup>0</sup>C) | 28.63±2.46<sup>a</sup> | 28.24±2.06<sup>a</sup> | 27.83±2.62<sup>a</sup> | 28.77±1.73<sup>a</sup> | 27.96±1.50<sup>a</sup> | 28.01±1.18<sup>a</sup> | 28.27±1.61<sup>a</sup> | 26.79±1.80<sup>a</sup> | 26.01±2.10<sup>a</sup> |
| Hydrogen Potential (pH) | 7.20±1.37<sup>c</sup> | 7.98±0.73<sup>c</sup> | 7.06±1.05<sup>c</sup> | 6.27±0.59<sup>b</sup> | 6.49±0.95<sup>b</sup> | 5.92±0.27<sup>a</sup> | 6.44±1.74<sup>b</sup> | 6.13±0.52<sup>b</sup> | 5.82±0.28<sup>a</sup> |
| Conductivity (ΨSCM<sup>-1</sup>) | 48.09±24.23<sup>d</sup> | 52.29±28.76<sup>d</sup> | 30.99±18.42<sup>c</sup> | 18.57±9.24<sup>a</sup> | 20.96±9.27<sup>a</sup> | 36.88±25.03<sup>c</sup> | 27.48±14.18<sup>b</sup> | 26.93±11.24<sup>b</sup> | 24.22±7.61<sup>a</sup> |
| Turbidity (NTU) | 7.41±2.97<sup>a</sup> | 7.36±6.50<sup>a</sup> | 42.43±14.40<sup>d</sup> | 31.43±11.72<sup>c</sup> | 11.14±5.05<sup>b</sup> | 3.35±1.18<sup>a</sup> | 48.24±20.57<sup>d</sup> | 4.99±1.56<sup>a</sup> | 1.78±0.66<sup>a</sup> |
| Dissolved Oxygen (DO) (mg/l) | 17.27±0.81<sup>a</sup> | 17.21±1.38<sup>a</sup> | 17.84±1.29<sup>a</sup> | 16.91±1.19<sup>a</sup> | 16.37±0.43<sup>a</sup> | 16.42±0.45<sup>a</sup> | 17.01±0.89<sup>a</sup> | 16.99±0.73<sup>a</sup> | 16.29±0.37<sup>a</sup> |
| (BOD) (mg/l) | 5.35±0.29<sup>a</sup> | 5.31±0.44<sup>a</sup> | 5.05±0.10<sup>a</sup> | 5.48±0.37<sup>a</sup> | 5.21±0.40<sup>a</sup> | 5.42±0.44<sup>a</sup> | 5.26±0.21<sup>a</sup> | 5.47±0.39<sup>a</sup> | 4.98±0.10<sup>a</sup> |
| (COD) (mg/l) | 40.06±12.15<sup>a</sup> | 37.27±8.79<sup>a</sup> | 39.61±11.84<sup>a</sup> | 41.78±13.07<sup>a</sup> | 39.72±6.80<sup>a</sup> | 32.03±9.65<sup>a</sup> | 33.50±7.15<sup>a</sup> | 32.75±6.61<sup>a</sup> | 32.64±6.57<sup>a</sup> |
| Acidity (mg/l) | 90.11±48.55<sup>a</sup> | 90.96±45.54<sup>a</sup> | 101.18±46.68<sup>a</sup> | 90.01±42.52<sup>a</sup> | 87.11±42.97<sup>a</sup> | 89.92±42.26<sup>a</sup> | 99.56±46.44<sup>a</sup> | 86.18±35.99<sup>a</sup> | 85.53±45.14<sup>a</sup> |
| Alkalinity (mg/l) | 103.73±60.46<sup>b</sup> | 119.56±58.6<sup>b</sup> | 18.17±7.42<sup>a</sup> | 134.96±50.03<sup>b</sup> | 119.23±65.28<sup>b</sup> | 119.15±65.15<sup>b</sup> | 130.93±43.65<sup>b</sup> | 134.76±47.90<sup>b</sup> | 117.73±63.97<sup>b</sup> |
| (TH) (mg/l) | 41.06±2.27<sup>a</sup> | 41.49±4.18<sup>a</sup> | 42.22±2.26<sup>a</sup> | 37.71±9.49<sup>a</sup> | 35.02±7.45<sup>a</sup> | 37.64±9.47<sup>a</sup> | 39.48±3.08<sup>a</sup> | 40.30±2.58<sup>a</sup> | 37.64±4.64<sup>a</sup> |
| TDS (mg/l) | 11.12±3.77<sup>a</sup> | 11.50±3.89<sup>a</sup> | 8.24±3.98<sup>a</sup> | 9.31±4.62<sup>a</sup> | 9.27±4.71<sup>a</sup> | 10.11±5.14<sup>a</sup> | 8.06±4.50<sup>a</sup> | 7.88±4.27<sup>a</sup> | 10.61±2.06<sup>a</sup> |
| TSS (mg/l) | 39.80±3.66<sup>b</sup> | 34.72±2.65<sup>a</sup> | 35.96±1.95<sup>a</sup> | 37.49±2.30<sup>b</sup> | 37.28±1.85<sup>b</sup> | 37.11±1.91<sup>b</sup> | 38.40±3.05<sup>b</sup> | 38.73±3.12<sup>b</sup> | 34.00±2.47<sup>a</sup> |
| Salinity (mg/l) | 0.12±0.08<sup>a</sup> | 0.14±0.10<sup>a</sup> | 11.50±10.60<sup>c</sup> | 15.74±11.29<sup>c</sup> | 0.07±0.01<sup>a</sup> | 1.71±0.76<sup>b</sup> | 25.71±5.35<sup>c</sup> | 0.09±0.08<sup>a</sup> | 0.01±0.02<sup>a</sup> |
| Chloride (mg/l) | 30.61±2.11<sup>b</sup> | 31.43±1.50<sup>b</sup> | 32.16±1.81<sup>b</sup> | 32.10±0.51<sup>b</sup> | 31.66±0.24<sup>b</sup> | 31.13±0.70<sup>b</sup> | 28.33±1.96<sup>a</sup> | 29.05±2.34<sup>a</sup> | 28.97±2.31<sup>a</sup> |
| Fluoride (mg/l) | 0.00±0.00<sup>a</sup> | 0.00±0.00<sup>a</sup> | 0.00±0.00<sup>a</sup> | 0.29±0.27<sup>b</sup> | 0.53±0.43<sup>b</sup> | 0.82±0.25<sup>c</sup> | 0.85±0.34<sup>c</sup> | 0.79±0.31<sup>a</sup> | 0.00±0.00<sup>a</sup> |
| Aluminum (mg/l) | 0.00±0.00<sup>a</sup> | 0.00±0.00<sup>a</sup> | 0.01±0.01<sup>a</sup> | 0.01±0.01<sup>a</sup> | 0.02±0.00<sup>b</sup> | 0.02±0.01<sup>b</sup> | 0.03±0.01<sup>b</sup> | 0.02±0.01<sup>b</sup> | 0.00±0.00<sup>a</sup> |
| Sodium (mg/l) | 12.32±2.38<sup>a</sup> | 16.06±1.99 | 12.22±2.72<sup>a</sup> | 14.54±0.80<sup>b</sup> | 14.45±1.07<sup>b</sup> | 14.65±0.90<sup>b</sup> | 16.39±0.20<sup>b</sup> | 15.48±2.40<sup>b</sup> | 12.85±2.63<sup>a</sup> |
| Potassium (K) (mg/l) | 2.65±0.76<sup>a</sup> | 2.49±0.61<sup>a</sup> | 3.14±0.03<sup>a</sup> | 3.23±0.56<sup>a</sup> | 2.97±0.64<sup>a</sup> | 3.07±0.52<sup>a</sup> | 3.10±0.04<sup>a</sup> | 3.29±0.17<sup>a</sup> | 2.42±0.59<sup>a</sup> |
| Calcium (Ca) (mg/l) | 52.22±7.67<sup>a</sup> | 52.58±8.57<sup>a</sup> | 56.20±8.18<sup>b</sup> | 59.76±6.15<sup>b</sup> | 59.72±6.47<sup>b</sup> | 50.31±6.60<sup>a</sup> | 59.98±6.29<sup>a</sup> | 51.99±7.61<sup>a</sup> | 50.47±6.77<sup>a</sup> |
| Magnesium (mg/l) | 137.23±11.45<sup>a</sup> | 136.11±12.02<sup>a</sup> | 132.71±48.02<sup>a</sup> | 129.99±37.38<sup>a</sup> | 129.26±37.07<sup>a</sup> | 129.66±37.10<sup>a</sup> | 146.67±21.78<sup>a</sup> | 145.74±19.58<sup>a</sup> | 129.26±37.07<sup>a</sup> |
| Iron (mg/l) | 2.23±0.42<sup>a</sup> | 2.29±0.73<sup>a</sup> | 3.27±0.98<sup>b</sup> | 2.96±2.18<sup>b</sup> | 4.01±0.12<sup>c</sup> | 2.06±0.02<sup>a</sup> | 5.16±1.74<sup>c</sup> | 3.37±1.60<sup>b</sup> | 1.21±0.20<sup>a</sup> |
| Zinc (mg/l) | 0.67±0.16<sup>a</sup> | 0.72±0.15<sup>a</sup> | 0.77±0.09<sup>a</sup> | 0.66±0.24<sup>a</sup> | 0.67±0.23<sup>a</sup> | 0.64±0.17<sup>a</sup> | 0.67±0.23<sup>a</sup> | 0.57±0.04<sup>a</sup> | 0.56±0.04<sup>a</sup> |
| Manganese (mg/l) | 0.02±0.01<sup>a</sup> | 0.03±0.02<sup>a</sup> | 0.02±0.02<sup>a</sup> | 0.03±0.02<sup>a</sup> | 0.04±0.02<sup>a</sup> | 0.03±0.02<sup>a</sup> | 0.02±0.02<sup>a</sup> | 0.03±0.03<sup>a</sup> | 0.00±0.00<sup>a</sup> |
| Cadmium (mg/l) | 0.01±0.02<sup>a</sup> | 0.02±0.02<sup>a</sup> | 0.00±0.00<sup>a</sup> | 0.01±0.03<sup>a</sup> | 0.00±0.00<sup>a</sup> | 0.00±0.00<sup>a</sup> | 0.02±0.03<sup>a</sup> | 0.02±0.03<sup>a</sup> | 0.00±0.00<sup>a</sup> |
| Lead (mg/l) | 0.12±0.04<sup>a</sup> | 0.11±0.07<sup>a</sup> | 0.09±0.08<sup>a</sup> | 0.11±0.05<sup>a</sup> | 0.09±0.06<sup>a</sup> | 0.13±0.07<sup>a</sup> | 0.14±0.06<sup>a</sup> | 0.12±0.08<sup>a</sup> | 0.11±0.04<sup>a</sup> |
| Copper (mg/l) | 0.03±0.03<sup>a</sup> | 0.04±0.03<sup>a</sup> | 0.03±0.03<sup>a</sup> | 0.04±0.04<sup>a</sup> | 0.03±0.04<sup>a</sup> | 0.05±0.04<sup>a</sup> | 0.03±0.04<sup>a</sup> | 0.03±0.04<sup>a</sup> | 0.03±0.03<sup>a</sup> |
| Chromium (mg/l) | 0.76±1.21<sup>a</sup> | 0.60±0.28<sup>a</sup> | 0.60±0.28<sup>a</sup> | 0.56±0.32<sup>a</sup> | 1.29±1.05<sup>a</sup> | 1.17±0.86<sup>a</sup> | 0.81±1.06<sup>a</sup> | 1.27±0.99<sup>a</sup> | 0.82±1.30<sup>a</sup> |
| Sulphate (mg/l) | 0.87±0.20<sup>a</sup> | 0.86±0.17<sup>a</sup> | 0.86±0.17<sup>a</sup> | 0.99±0.22<sup>b</sup> | 0.92±0.02<sup>b</sup> | 0.85±0.04<sup>a</sup> | 0.84±0.05<sup>a</sup> | 0.85±0.11<sup>a</sup> | 0.94±0.12<sup>b</sup> |
| Ammonia (mg/l) | 2.63±1.01<sup>a</sup> | 2.79±1.11<sup>a</sup> | 2.79±1.11<sup>a</sup> | 2.79±1.20<sup>a</sup> | 2.76±1.08<sup>a</sup> | 2.80±1.10<sup>a</sup> | 2.75±1.06<sup>a</sup> | 2.22±1.00<sup>a</sup> | 2.38±1.03<sup>a</sup> |
| Phosphate (mg/l) | 0.15±0.16<sup>a</sup> | 0.38±0.02<sup>a</sup> | 0.38±0.02<sup>a</sup> | 0.18±0.15<sup>a</sup> | 0.24±0.14<sup>a</sup> | 0.21±0.14<sup>a</sup> | 0.20±0.13<sup>a</sup> | 0.23±0.20<sup>a</sup> | 0.24±0.23<sup>a</sup> |
| Nitrite (mg/l) | 1.90±1.12<sup>a</sup> | 1.70±1.06<sup>a</sup> | 1.70±1.06<sup>a</sup> | 2.33±1.60<sup>a</sup> | 2.10±1.62<sup>a</sup> | 1.66±1.08<sup>a</sup> | 1.90±1.09<sup>a</sup> | 1.95±1.15<sup>a</sup> | 1.61±0.48<sup>a</sup> |
| Nitrate (mg/l) | 2.87±1.28<sup>a</sup> | 2.34±0.80<sup>a</sup> | 2.34±0.80<sup>a</sup> | 3.23±0.71<sup>b</sup> | 3.36±1.11<sup>b</sup> | 2.33±0.76<sup>a</sup> | 1.90±0.17<sup>a</sup> | 2.23±0.67<sup>a</sup> | 2.01±0.16<sup>a</sup> |
| Nickel (mg/l) | 0.97±0.61<sup>a</sup> | 0.91±0.26<sup>a</sup> | 0.91±0.26<sup>a</sup> | 1.00±0.42<sup>a</sup> | 0.90±0.25<sup>a</sup> | 0.84±0.22<sup>a</sup> | 0.94±0.15<sup>a</sup> | 0.94±0.17<sup>a</sup> | 0.95±0.15<sup>a</sup> |
| TPH (mg/l) | 14.86±0.38<sup>d</sup> | 10.41±4.55<sup>c</sup> | 13.861±1.21<sup>d</sup> | 13.00±1.52<sup>d</sup> | 2.81±1.58<sup>b</sup> | 11.57±1.62<sup>c</sup> | 4.07±0.94<sup>b</sup> | 3.84±0.45<sup>b</sup> | 0.001±0.00<sup>a</sup> |

**Table 4:**
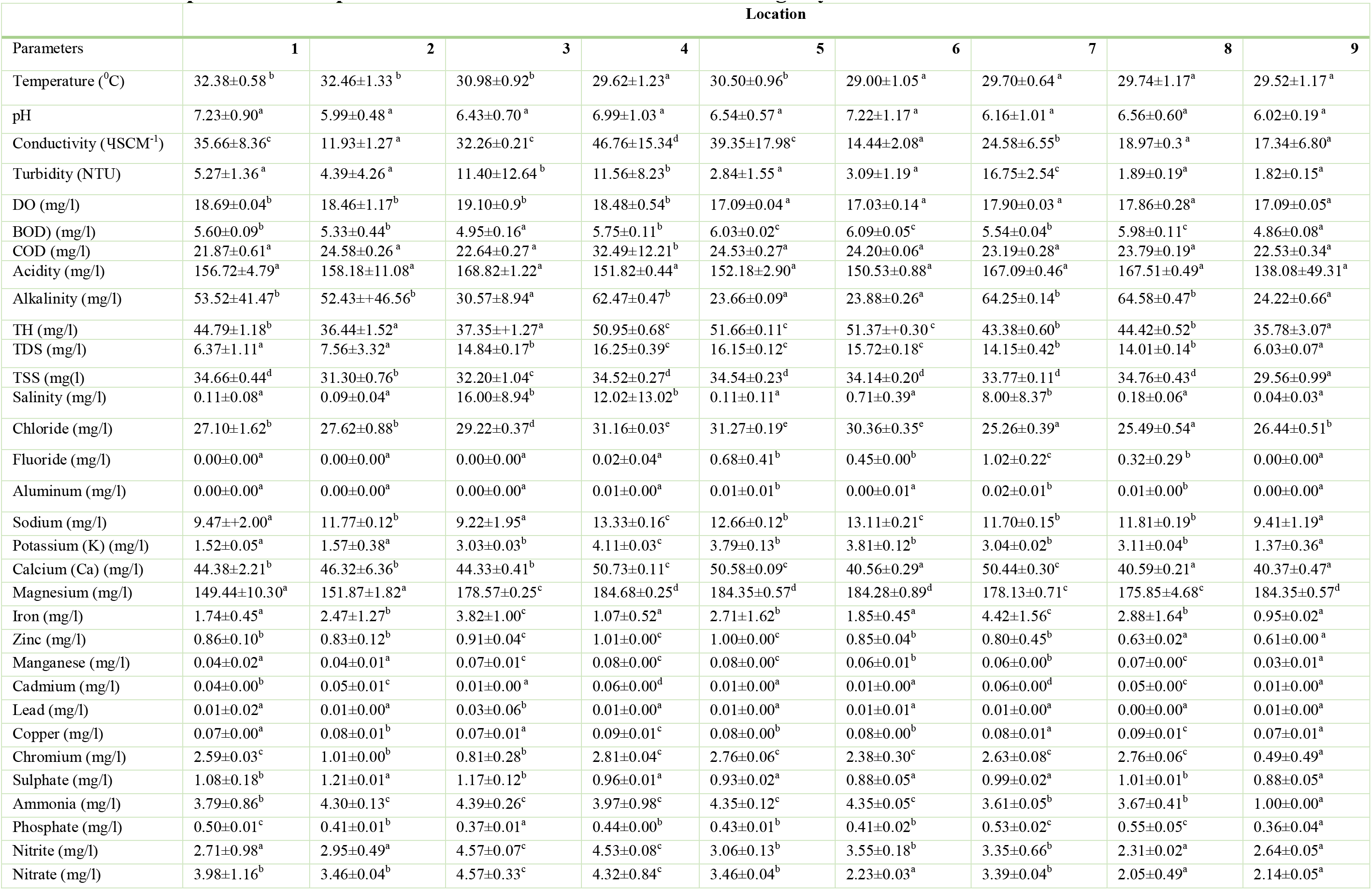
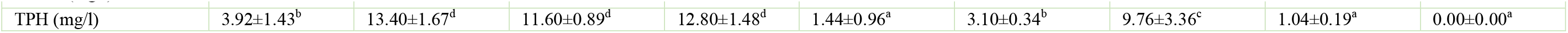
Comparison of the parameters in the different locations during dry season.

## 6. Discussion

### 6.1 Compare differences in water quality parameters in the study area (determine the level of pollutions in the different sites)

The phrase “trace elements” refers to a class of ubiquitous elements that are common in the environment, yet are harmful to people and organisms at very low quantities. Heavy metals (metals having a large atomic mass) for instance chromium (Cr), cadmium (Cd), lead (Pb), nickel (Ni), cobalt (Co), copper (Cu), mercury (Hg), tin (Sn), as well as zinc (Zn). While non-metals that remain viewed as trace elements comprise antimony (Sb), arsenic (As), as well as selenium (Se). Trace elements remain long-lasting and are not destroyed by metabolic activities. Trace elements exist in a variety of forms, including oxides, salts, organometallic complexes, sulphides, and dissolved ions in groundwater as well as soil solution. Chemical processes are driven by the partitioning of water, air as well as soil through particles adsorption or pH-dependent water dissolution (Alloway, 2012; Olalekan *et al.*, 2018b; Olalekan *et al.*, 2020a; Morufu *et al.*, 2021a). Trace elements remain geogenic (natural) in origin, because several rocks comprise significant quantities of trace elements that remain released into the milieu via anthropogenic or weathering action. Thus, tables 3&4 provide the statistical analysis findings for the physicochemical parameters. The Ebocha-Obrikom area of Rivers State is significant for irrigation, drinking, as well as industrial uses (Raimi and Sabinus, 2017b; Morufu and Clinton, 2017; Olalekan *et al.*, 2018b; Olalekan *et al.*, 2020a). Over the previous three decades, the Ebocha-Obrikom area of Rivers State has been significantly altered by population expansion as well as increased agricultural productivity. However, various small-scale entrepreneur activities and anthropogenic activities such as extractive industries and it related activities, as well as chemicals overuse in agriculture remain hypothesized toward degrading groundwater quality in Ebocha-Obrikom area of Rivers State and toxic element contamination of the food chain. The groundwater investigation of toxic materials in the Ebocha-Obrikom area of Rivers State is consequently vital for human health protection. A detailed analysis of groundwater geochemistry as well as associated estimation of community’s health risk that are visible to the groundwater, remain yet to be carried out, despite the fact that a clear understanding of the utmost significant aspects regulating the health risks is vital towards taking effective management measures for the residents regarding drinking water. Thus, the concentrations of analysed water quality parameters is summarized in Table 3&4. A total of thirty-four (34) water quality parameters were analysed during raining and dry season respectively. Eighteen (18) parameters such as temperature, pH, conductivity, turbidity, DO, BOD, Acidity, TSS, Salinity, Fluoride, Aluminum, Potassium, Magnesium, Iron, Zinc, Manganese, Cadmium and TPH were lowest at location nine (9) during the raining season. For dry season, twenty-two (22) parameters, which include: turbidity, BOD, Acidity, TH, TDS, TSS, Salinity, Fluoride, Aluminium, Potassium, Calcium, Iron, Zinc, Manganese, Cadmium, Copper, Chromium, Sulphate, Ammonia, Phosphate, Nickel and TPH recorded minimum values below limits of detection in sampling locations nine (9). Thus, the results showed a significant disparity between the various sampling locations. As it is evident that samples from location 1 to location 9 must remain adequately observed since, there may be a remarkable increase in these heavy metal level in the future, which could eventually cause health-related threats for indigenous residents. The temperature is one of the utmost critical elements influencing the biological activity of an aquatic organisms. Discern that there is high temperature variation in this region. Temperature was found to be highest with an average value of (28.77 – 32.46)^0^C at location 4 and 2 during the raining and dry seasons. While it was lowest (26.01 – 29.00) ^0^C at location 9 and 6 during the raining and dry season. The maximum permissible limit for temperature has not been stated but ambient in nature. Groundwater temperature tend to be influenced more by dry season than raining season. Thus, Morufu and Clinton (2017); Raimi and Sabinus, (2017b) and Olalekan *et al.*, (2018b) indicated that locations predisposed to the release of industrial pollutants and gas flaring typically have temperatures that are higher than those of their surrounding environments. Unarguably, the gas flare site must have influenced an increase in air temperature, thus correspondingly increasing the temperature of groundwater. This is indicative of groundwater pollution since anthropogenic activities seem to have influence groundwater pollution more in the dry season. Likewise, rising temperature may adversely impact agriculture, thereby increasing the vulnerability of marginalized agriculture-dependent rural populations of “Ebocha-Obrikom Community in Ogba/Egbema/Ndoni Local Government Area of Rivers State, which is home to Agip’s Ebocha, Obrikom as well as Obiafu Oil and Gas Facilities”. Thus, gas flare site can be a catalyst for such migrants to relocate in search of better living conditions and alternate livelihoods. Despite, the fact that the community is abundant in natural gas as well as is home to the state-owned gas turbine facility. There are several large and functioning gas stacks in the community. The people of Ogba remain primarily fishermen as well as farmers who rely on groundwater and small streams particularly the Orashi River as community water source. Water pH represents hydrogen ion concentration as well as is affected by the interaction of several compounds dissolved in water, aquatic animal respiration, photosynthetic activity of aquatic plants, as well as organic matter decomposition. The natural water pH influences biological as well as chemical interactions, and controls the metal ions solubility as well as the consequences on natural aquatic life. The pH range in which aquatic creatures thrive varies (Olalekan *et al.*, 2018b; Olalekan *et al.*, 2020a). It is in itself poisonous at a certain level as well as been shown toward influencing the toxicity of heavy metals, cyanides, hydrogen sulfide as well as ammonia. Even though the pH value of water still remains a significant indicator of its acidity or alkalinity. Water hydrogen ion has no direct detrimental influence on consumers, as well as an indirect outcome, but it can cause changes in several characteristics of water quality, such as the primary chemical formation as well as survival of infectious microorganisms in water. Aside from triggering irritation of the digestive tract in persons with high sensitivity (Olalekan *et al.*, 2018b; Ukah *et al.*, 2019; Olalekan *et al.*, 2020a; Morufu *et al.*, 2021a). The pH has no direct impact on human heath, it can change water taste as well as exhibit linked to some other water quality characteristics. The carbonate cycle composed of CO_2_, H_2_CO_3_, HCO_3_ and CO_3_ , which plays a role for regulating pH. The importance of alkalinity lies in its role for carbon dioxide chemistry, trace metal speciation and buffer capacity of the groundwater. Thus, hydrogen-ion-concentration (pH) is a master control measure in the environment that demonstrates the chemical as well as biological features of water as well as it is used in identifying waters alkalinity or acidity. It is remarkably critical to understand the water nature, as well as it also reveals a close relation with other water chemical constituents. The main variables affecting the variation of pH in any milieu are: dissolved oxygen, water temperature, land runoff, decomposition of organic matter photosynthetic activity respiration of aquatic organisms and some physic-chemical processes, such as precipitation and oxidation reduction tacking place in the environment. All of the aquifers in the raining season contained groundwater that had slightly acidic to slightly alkaline nature of groundwater, with on-site pH values ranging from 5.82 to 7.98. The highest pH was found in location 2, possibly due to more intensified human activity in such location and the lowest was detected in location 9, indicating that gas flaring at Ebocha-Obrikom area of Rivers State seems to have affected groundwater acidity. Albeit the waters may not show the apparent threat to human health dependent on pH, they would be destructive to picturesqueness after some time (Morufu and Clinton, 2017; Raimi and Sabinus, 2017b; Olalekan *et al.*, 2018b; Egbueri *et al.*, 2019; Olalekan *et al.*, 2020a; Morufu *et al.*, 2021a). Similarly, the analytical results during the dry season discovered that pH ranged from 5.99 to 7.23, which was within the WHO (2017) acceptable pH range of 6.5 to 8.5 for drinking water, except at sampling location 1, which had the highest pH value. Thus, on pH scale, a number of 7 denotes neutral water, a value less than 7 denotes acidic water, and any value greater than 7 denotes basic water. Hence, the increasing pH values could lead to increase in the rate of corrosion. In any of the groundwater tests, nonetheless, no location was determined to remain beyond the maximum permissible limit. The pH ground water variation in Ebocha-Obrikom area was below the permitted level and was therefore, not hazardous for drinking (Olalekan *et al.*, 2018b; Olalekan *et al.*, 2020a; Morufu *et al.*, 2021a). For electric conductivity (EC), it is a measure of the ability of ions in a solution to carry electric current (Olalekan *et al.*, 2018b; Afolabi and Raimi, 2021; Morufu *et al.*, 2021a). This ability depends on the presence of ions, their total concentration, mobility, valence and relative concentration and temperature. Warmer the water, higher the conductivity. Conductivity in water is affected by the presence of inorganic dissolved solids such as chloride, nitrate, sulfate and phosphate anion or sodium, magnesium, calcium, iron and aluminum cautions. Organic compounds like oil, phenol, alcohol and sugar do not conduct electrical current very well and therefore have low conductivity when in water. According to Kanga *et al.*, (2020), the EC is often used to calculate the ionic concentration of groundwater, which fluctuates based on the concentration, ions type presents in water, as well as temperature. The most important test that reveals the total concentration of soluble salts is the conductivity test. Thus, electrical conductivity (EC) diverges from (24.22 - 52.29)(11.93 – 46.76)μs/cm, with an average of (31.83 – 26.81)μs/cm (Table 3&4). Because it is a measure of a material’s capacity toward conducting an electric current, the disparity suggests a wide range of salts existing in groundwater. TDS is indicated in terms of the degree of salt concentration in the studied area. Thus, the percolation of agrochemicals and natural groundwater recharge processes increases the EC value (Ako *et al.*, 2014; Olalekan *et al.*, 2018b; Afolabi and Raimi, 2021; Morufu *et al.*, 2021a). Hence, higher values for conductivity at location 2 &4 could be attributed to excessive accumulation of dissolved salts, spilled oil through possible emission of flared gases getting into groundwater, agricultural lands and other organic materials present as natural resources. Equally, salinization could affect quality of groundwater in the research study area in the following years. Although, samples from gas flared environment are relatively more conductive. Locally, the effect of salt water infiltration into the aquifer from the tide influenced by river Orashi may also be an important factor in the salinization of groundwater in the area (Olobaniyi and Owoyemi, 2006; Raimi and Sabinus, 2017b; Olalekan *et al.*, 2018b; Afolabi and Raimi, 2021; Morufu *et al.*, 2021a). The result found support in Ehirim and Nwankwo, (2010); Morufu and Clinton, (2017); Olalekan *et al.*, (2018b) and Morufu *et al.*, (2021a) which established that electrical conductivity values of the ground water samples collected from the studied location are observed to be low throughout the sampling locations and the variations of their mean concentrations at different distances. According to Okafor and Opuene (2007); Morufu and Clinton, (2017) and Olalekan *et al.*, (2018b) the electrical conductivity indicates the level of salinity; hence it greatly affects water taste as well as represents a remarkable impact on the user’s acceptance. For turbidity, the American Public Health Association (APHA, 2012) and Morufu and Clinton, (2017) defines turbidity as “the optical property of a water sample that causes light to be scattered and absorbed rather than transmitted in straight lines through the sample”. In simple terms, turbidity answers the question, “How cloudy is the water?” The ability of light to pass through water is directly proportional to the volume of suspended particles within the water body. The higher the volume of suspended particles, the cloudier the water becomes. Turbidity is measured using an electronic turbidity meter. The results are reported in Nephelometric Turbidity Units (NTU) or by filtering a water sample and comparing the filter’s colour (how light or dark it is) to a standard turbidity chart. APHA specifies drinking water turbidity shall not exceed 5 NTUs. Thus, turbidity conditions may increase the possibility for waterborne disease (Raimi *et al.*, 2017; Olalekan *et al.*, 2018b; Olalekan *et al.*, 2020a; Afolabi and Raimi, 2021; Morufu *et al.*, 2021a). If turbidity is largely due to organic particles, dissolved oxygen depletion may occur in the water body (Morufu and Clinton, 2017; Morufu *et al.*, 2021a). Higher turbidity levels are often associated with higher levels of disease-causing microorganisms such as viruses, parasites and some bacteria (Raimi *et al.*, 2017; Olalekan *et al.*, 2019a; Raimi *et al.*, 2019a). These organisms can cause symptoms such as nausea, cramps, diarrhea, and associated headaches (USEPA, 2015; Raimi *et al.*, 2017; Olalekan *et al.*, 2020a; Afolabi and Raimi, 2021; Morufu *et al.*, 2021a). Thus, highest value of turbidity was noticed at location 7 (48.24 – 16.75) NTU during the raining and dry season. Groundwater turbidities remained below the usual recommended maximum tolerable limit of 5NTU for drinking water at location 6 & 9 for raining season and location 5, 6, 8 & 9 for dry seasons. Although, location 8 (raining season) and location 1 & 2 (dry seasons) approximately attained the maximum permissible limit for drinking water. Thus, turbidities were higher during raining season than dry season. Hence, wet season influence turbidity more than its counterpart dry seasons. This could be due to continuous and impactful predisposition to receiving large quantities of organic and inorganic material emanating from gas flaring and oil spillage contaminating the ground water of the study area. Dissolved Oxygen studies in water are very important since it is considered one of the most important limiting factors for the life of aquatic organisms which affect the biological processes of the aquatic organisms, the respiration of animal and oxidation of the organic matter in water and sediments. It is an important parameter in assessing the degree of pollution in which sewage pollution has been generally regarded as an organic pollution affecting fish and other aquatic life, principally through oxygen depletion. Bacteria consume oxygen as organic matter decay. As a result, an oxygen deficient environment can develop in lakes and rivers with the excess organic matter. These conditions can eventually lead to fish kills and can result in reduced growth, disruption of life cycles, migration to avoid poor condition and death of benthic organisms (Amiri *et al.*, 2014; Amiri *et al.*, 2015; Morufu and Clinton, 2017; Raimi and Sabinus, 2017b; Olalekan *et al.*, 2018b; Olalekan *et al.*, 2020a; Afolabi and Raimi, 2021; Morufu *et al.*, 2021a). Thus, highest value of Dissolved Oxygen (DO) for groundwater was noticed at location 3 (17.84 – 19.10) mg/L for both raining and dry seasons. It was observed that DO was higher during both seasons. Thus, indicating higher biological activity due to higher rate of anthropogenic activities. Meanwhile, studies by Chapman and Kimstach (1992); Morufu and Clinton, (2017); Raimi and Sabinus, (2017b); Olalekan *et al.*, (2018b) and Morufu *et al.*, (2021a) found that DO concentration below 5mg/L have a negative impact on the functioning as well as biological communities’ survival and concentrations below 2mg/L may result in the death of more life. The BOD is the amount of oxygen consumed by bacteria in the decomposition of organic material. It also includes the oxygen required for the oxidation of various chemical in the water such as sulfides, ferrous iron and ammonia (Amiri *et al.*, 2015; Morufu and Clinton, 2017; Raimi and Sabinus, 2017b; Olalekan *et al.*, 2018b; Morufu *et al.*, 2021a). This parameter provides information on the potential for microbial respiration to breakdown the organic material present in the water which can lead to low DO and is a suggested cause of hypoxia (Morufu and Clinton, 2017; Raimi and Sabinus, 2017b; Olalekan *et al.*, 2018b; Morufu *et al.*, 2021a). Even though, biological oxygen demand (BOD) reflects the amount of oxygen needed by bacteria. It is used to determine any receiver environment pollution potential as well as assimilation capacity. The present study for BOD had its highest value at location 4 & 6 (5.48 – 6.09) mg/L during raining and dry seasons. Dry season had higher values than raining season. In consequence, it might be extrapolated that anthropogenic activities may influenced increased BOD during the dry season, while supporting higher biochemical activity. However, irrespective of seasonal variations, both seasons influenced BOD. Hence, gas flaring must have contributed to this trend. For the chemical oxygen demand (COD), it is the amount of oxygen required to oxidize the organic matter in waste water by use of a strong oxidant and to convert it to carbon dioxide and water. (Odipe *et al.*, 2018; Henry *et al.*, 2019a; Afolabi and Raimi, 2021; Morufu *et al.*, 2021a). COD test is used to assess the degree of pollution in the area under investigation. The value of COD is always higher than that of BOD_5_ because many organic substances can be oxidized chemically but not biologically (Odipe *et al.*, 2018; Henry *et al.*, 2019b; Afolabi and Raimi, 2021; Morufu *et al.*, 2021a). Thus, chemical oxygen demand (COD) is used in defining the level of pollution in water. Once the level of COD in the water exceeds 25 mg/L, it shows that there is higher concentration of pollutants. Furthermore, if the levels of COD exceed 50 mg/L, it is indicative that there is severe pollution which can be toxic for aquatic life. COD values were found to be highest at location 4 (41.78 – 32.49) mg/L during raining and dry season. This indicates that organic pollution of water is more severe during raining season than dry seasons. Thus, convincing the statement that organic pollution is worse at raining season is more credible. All reported values in this investigation were above the maximum acceptable limit of 10mg/L for COD, as the trend shows higher chemical activity and could be attributed to areas prone to gas flaring and oil spill related activities impact. Since COD measures oxygen demand created by organic and inorganic compounds (Odipe *et al.*, 2018; Henry *et al.*, 2019b; Afolabi and Raimi, 2021; Morufu *et al.*, 2021a). For acidity, this arises from the presence of weak or strong acids and/or certain inorganic salts. The presence of dissolved carbon dioxide is usually the main acidity factor in unpolluted surface and ground waters. There is no particular implication apart from palatability consideration in excessively acid waters (USEPA, 2015; Morufu *et al.*, 2021a). The acidity of water will affect its corrosiveness and also the speciation of some of its other constituents. Thus, acidity values range from highest at location 3 (101.18 – 168.82) mg/L for both raining season and dry season (Table 3&4). There is currently no maximum value set for acidity according to WHO/SON/NAFDAC standards of potability. Also, the alkalinity of a natural body of water is generally due to the presence of bicarbonates formed in reactions in the soil through which water percolates. It is a measure of the capacity of water to neutralize acids and it reflects its so-called buffer capacity (USEPA, 2015; Morufu and Clinton, 2017; Raimi and Sabinus, 2017b; Olalekan *et al.*, 2018b; Olalekan *et al.*, 2020a; Afolabi and Raimi, 2021). Alkalinity in natural waters may be attributable to bicarbonate and hydroxides. Alkalinity is involved in the consequential effects of eutrophication of water (USEPA, 2015; Morufu and Clinton, 2017; Raimi and Sabinus, 2017b; Olalekan *et al.*, 2018b; Olalekan *et al.*, 2020a; Afolabi and Raimi, 2021). Alkalinity was highest at location 4 & 8 (134.96-64.58) during the raining and dry season and lowest at location 3 & 5 (18.17 – 23.66) during raining and dry season correspondingly. The high rate might remain attributed toward constant discharge of acidic and chemicalized substances through oil spillage and gas flaring which latter find their ways into the groundwater bodies and adjoining environment. Traditionally, for hardness, it is the measure of water’s ability toward reacting with soap as well as characterizes water’s ability toward binding soap to form scum or lather, which is a reaction that is chemically harmful toward the process of washing (Morufu and Clinton, 2017; Raimi and Sabinus, 2017b; Olalekan *et al.*, 2018b; Olalekan *et al.*, 2020a; Afolabi and Raimi, 2021; Morufu *et al.*, 2021a). Hardness of water is caused by groundwater interacting with rock formations. It is the sum of the concentrations of dissolved polyvalent metal ions which Ca^2+^ and Mg^2+^ are predominant. Calcium hardness, regardless of the salts involved, is referred to as calcium hardness. Similarly, magnesium hardness refers to hardness caused by magnesium. Because calcium as well as magnesium remain the only remarkable minerals that are known to induce hardness. The sources of the metallic ions remain often found in sedimentary rocks, the most prevalent of which remain limestone (CaCO_3_) as well as dolomite (CaMg(CO_3_)_2_) (Morufu and Clinton, 2017; Raimi and Sabinus, 2017b; Olalekan *et al.*, 2018b; Olalekan *et al.*, 2020a; Afolabi and Raimi, 2021; Morufu *et al.*, 2021a). Total Hardness (TH) as calcium carbonate, is the sum of Ca^2+^ as well as Mg^2+^ in groundwater and signifies the existence of alkaline earths. Hardness can induce encrustation of water supply distribution systems (Morufu and Clinton, 2017; Raimi and Sabinus, 2017b; Olalekan *et al.*, 2018b; Mahato *et al.*, 2018; Olalekan *et al.*, 2020a; Afolabi and Raimi, 2021; Morufu *et al.*, 2021a). The concentration of TH levels varied from 39.17mg/L to 44.02mg/L [both raining and dry season]. More than half of the sample’s groundwater in Ebocha-Obrikom area of Rivers State possess TH below 100mg/L. Thus, total hardness (TH) of the aquifers fluctuated on average from 39.17mg/L for raining season to 44.02 mg/L for dry seasons, with the lowest confined groundwater (mean = 35.02 at location 5) (35.78 at location 9), and highest confined groundwater (mean = 42.22 mg/L at location 3) (51.66 at location 5). TH represents the total concentration of Ca^2+^ as well as Mg^2+^ in this system, which was mostly caused by mineral dissolution like carbonates as well as gypsum (Li *et al.*, 2016c; Morufu and Clinton, 2017; Raimi and Sabinus, 2017b; Olalekan *et al.*, 2018b; Morufu *et al.*, 2021a). Koffi *et al*. (2017) discovered that the increased groundwater hardness was caused by carbonate sources. The research results contradicted Disli (2017)’s finding that TH level varied “from 198.5 to 409.5 mg/L, with a mean of 289.1 in the Upper Tigris River Basin, Diyarbakır–Batman, Turkey”. Furthermore, in the crystalline basement complex rock of India, Adimalla *et al*. (2018a) recorded TH values ranging from 60 to 750 mg/L, with a mean of 202 mg/L, and approximately 18% of the samples falling into the moderately hard category, whereas Koffi *et al*. (2017) obtained TH values ranging from 50.8 to 272 mg/L, with 60.6% of samples falling into the moderately hard category. Nonetheless, the highest permitted amount of TH aimed at drinking purposes is 500 mg/L, with a preferred limit of less than 100 mg/L (WHO, 2017). The groundwater in the Ebocha-Obrikom oil and gas area was found to be 100% safe, with all samples falling within the maximum permitted 500 mg/L limit. Conversely, Ezekwe *et al.*, (2012), Ukah *et al.*, (2019) claims that subsurface waters remain often tougher than surface waters. While, Total Dissolved Solids (TDS) on the other hand, refers to the various minerals that remain existent in water in dissolved form and is a pointer of water salinity as well as signifies dissolved salts in water, such as large calcium, chlorides, bicarbonates, phosphates, sulfates, carbonates, silica, potassium, magnesium and sodium (WHO, 2006; Morufu and Clinton, 2017; Raimi and Sabinus, 2017b; Olalekan *et al.*, 2018b; Morufu *et al.*, 2021a). TDS levels above a certain threshold impair the palatability of water as well as promote gastrointestinal discomfort in consumers (Raimi *et al.*, 2017; Olalekan *et al.*, 2020a; Morufu *et al.*, 2021a). Consuming water of high TDS for an extended period of time can result in kidney stones. Consequently, it’s a crucial factor to consider while assessing drinking water consistency as well as other water forms. It is also, an important metric for determining the appropriateness of irrigation as well as drinking water. WHO (2017) claims that groundwater taste with a TDS level of less than 600 mg/L is regarded good for aquatic lives, fisheries and residential water supply protection, but TDS levels greater than 1000 mg/L remain well-thought-out as unpalatable for the purposes of drinking? Thus, TDS levels beyond the WHO groundwater specified threshold may cause unpleasant taste as well as gastrointestinal complications (Morufu and Clinton, 2017; Raimi and Sabinus, 2017b; Olalekan *et al.*, 2018b; Morufu *et al.*, 2021a). High TDS maybe derived from intensive or massive usage of agrochemical, dissolution of salts, ion exchange, organic materials, and sediment dissolution, aquifer percolation and allied substances emanating from oil related activities such as gas flaring as well as oil spillage (Chabukdhara *et al.*, 2017; Morufu and Clinton, 2017; Raimi and Sabinus, 2017b; Olalekan *et al.*, 2018b).

Thus, groundwater contamination in this wise could be due to the continuous contamination of groundwater by industrial pollutants as suggested by Olalekan *et al.*, (2018b) and Olalekan *et al.*, (2020a). Although, all values were significantly below the acceptable limit, these total dissolved solids (TDS) show a very weak variability as seen by their low standard deviation (SD). Its concentration varies from 9.57mg/L to 12.34mg/L [both raining and dry season]. The overall hydro chemical groundwater characteristics are regulated by major ions (Morufu and Clinton, 2017; Raimi and Sabinus, 2017b; Olalekan *et al.*, 2018b; Li *et al.*, 2018c; Afolabi and Raimi, 2021; Morufu *et al.*, 2021a). Hence, the groundwater samples were desirable as well as allowed for purposes of drinking based on the TDS categorization of all collected groundwater samples. Thus, the difference in the concentration of TDS indicates a wide variation in the geochemical processes. Meanwhile, if the waters are to be utilized in fish ponds, WHO (2017) recommends a concentration of 1500mg/L for fisheries and aquatic life protection, and for household water supply. Because all values remained below the tolerable limit, they remain safe for drinking on TDS basis as supported by researches from Dami, Ayuba and Amukali (2012); Morufu and Clinton, (2017); Raimi and Sabinus, (2017b) and Olalekan *et al.*, (2018b) that waters within the limits attained here remain extremely palatable. Thus, all of the sample’s groundwater showed TDS levels that were lower than the WHO (2017) standard (250 mg/L) as well as the NAFDAC threshold levels (500 mg/L). Concentrations of TDS in Ebocha-Obrikom area remain below the optimal threshold in all locations, according to earlier research, Besides, TDS concentrations remained sufficient in quality for drinking in all areas (Olalekan *et al.*, 2018b; Morufu *et al.*, 2021a). According to Adimalla & Qian (2021), about 95% of the total samples remained below the ideal drinking threshold (TDS: 1000 mg/L), with the samples remaining appropriate for irrigation (TDS: 1000–3000 mg/L). For total suspended solids (TSS), the mean values show that the highest value in groundwater was observed at location 1&8 (39.80 – 34.76) mg/L for raining and dry seasons. While, the least value of (34.00 – 29.56) mg/L at location 9 for raining and dry seasons respectively. All of the values noted in this investigation were above the maximum allowable limit. Indicating that gas flaring and oil spillage releases persistent non-combustible chemicals and less dense volatile chemicals into the environment. In addition, it could be due to massive percolation of water through the water table and discharge of large quantities of substances directly into groundwater bodies or out rightly onto terrestrial regions where they leach into ground water bodies. For salinity, all groundwater contains salts in solution; reported salt contents range from less than 25mg/L in a quartzite spring to more than 300,000mg/L in brines (David, 2006). The type and concentration of salts depends on the environment, movement and source of the groundwater. Ordinarily, higher concentration of dissolved constituents is found in groundwater than in surface water because of the greater exposure to soluble materials in geologic strata. Soluble salts in groundwater originate primarily from solution of rock materials. Bicarbonate, usually the primary anion in groundwater, is derived from carbon dioxide released by organic decomposition in the soil. Salinity varies with specific surface area of aquifer materials, solubility of minerals and contact time; values tend to be highest where movement of groundwater is least; hence salinity generally increase with depth. Thus, salinity values range from highest at location 7 (25.71) mg/L for raining season to location 3 (16.00) mg/L (Table 3&4). The maximum value is set at 600 mg/L according to WHO/SON/NAFDAC standards of potability. All values recorded in this study were below the maximum permissible limit of 600 mg/l for drinking water. Chloride is found in many chemical and other substances in the body. It is an important part of the salt found in many foods and used in cooking. It is also an essential part of the digestive (stomach) juices. It is found in table salt or sea salt as sodium chloride. Too much chloride from salted foods can increase blood pressure, even in young children (Goldman *et al.*, 1985; Morufu and Clinton, 2017; Raimi and Sabinus, 2017b; Olalekan *et al.*, 2018b; Morufu *et al.*, 2021a) and cause a buildup of fluid in people with congestive heart failure, cirrhosis, or kidney disease (www.health.nytimes.com). Although excessive intake of drinking-water containing sodium chloride at concentrations above 2.5 g/litre has been reported to produce hypertension (Olalekan *et al.*, 2018b; Morufu *et al.*, 2021a), this effect is believed to be related to the sodium ion concentration. Thus, chloride is an inert as well as mobile compound, whose natural amount is determined by sea distance, geographical location as well as precipitation amount, but also by the regional impact of groundwater resulting from saline water inputs. A number of studies alleged that Cl excess in groundwater is an indicator pollution index and has a harmful influence on human health (Morufu and Clinton, 2017; Raimi and Sabinus, 2017b; Olalekan *et al.*, 2018b; Adimalla *et al.*, 2018a; Li *et al.*, 2018a; Morufu *et al.*, 2021a). Though, chloride is also one of the prominent anion in Rivers State oil and gas producing area of Ebocha-Obrokom, ranging from (28.33 – 32.16)(25.26 – 31.27) mg/L for both raining and dry season with a mean of (30.60 – 28.21) mg/L (Table 3&4). When compared to the acceptable limit of 200 mg/L, all of the samples had lower concentrations, which might be attributable to higher neutralization reactions by dissolved alkaline hydroxyl-containing substances. It has been noted that while water with low chloride ions is not dangerous, chloride ions at large concentrations can kill floras when used for horticultural or agricultural applications. It may also be to blame for the unpleasant taste of water consumed (WHO, 2004). While samples at location (3 & 5) for raining and dry season were high in the Ebocha - Obrikom region. These samples were collected at the center of the research area. Elevated levels of chloride may perhaps remain linked to domestic waste effluents, septic tanks leakage, as well as chloride bearing rocks dissolution (Koffi *et al.*, 2017; Morufu and Clinton, 2017; Raimi and Sabinus, 2017b; Olalekan *et al.*, 2018b; Adimalla and Li, 2018; Adimalla and Venkatayogi, 2018; Olalekan *et al.*, 2020a; Afolabi and Raimi, 2021; Morufu *et al.*, 2021a). Thus, the numerous natural as well as chlorides anthropogenic sources, concentrations of ambient background vary greatly. In spite of the fact that no health dangers have been established, residents of Ebocha-Obrikom areas remain hesitant toward drinking water due to texture as well as taste issues. Hence, high Cl^−^ groundwater concentrations remain seen as a symptom of pollution from a number of sources, and they impart a salty flavor to the water (Marghade *et al.*, 2012; Morufu and Clinton, 2017; Raimi and Sabinus, 2017b; Olalekan *et al.*, 2018b; Morufu *et al.*, 2021a). In addition, Chloride concentration in drinking water above 200 mg/l have been linked to heart disease, asthma as well as possibly cancer. For fluorine (F^−^), it is widely distributed in the environment. It is usually safe to drink water within the limits of 0.5–1.5 mg/l according to the suggested guidelines (WHO, 2017; Morufu and Clinton, 2017; Raimi and Sabinus, 2017b; Olalekan *et al.*, 2018b; Yousefi *et al.*, 2019a; Olalekan *et al.*, 2020a; Afolabi and Raimi, 2021; Morufu *et al.*, 2021a). Fluoride becomes harmful to health at quantities above/below this recommendation, and is denoted as a double-edged sword (Olalekan *et al.*, 2020a). Water consumers remain prone to dental carries at lower concentration, while at larger concentrations, it can induce skeletal fluorosis, debilitating fluorosis, dental fluorosis, as well as kidney damage (Morufu and Clinton, 2017; Raimi and Sabinus, 2017b; Olalekan *et al.*, 2018b; Olalekan *et al.*, 2020a; Morufu *et al.*, 2021a). Excess consumption of fluoride has also been associated to infertility, abortion, fertility, as well as hypertension (Yousefi *et al.*, 2018, 2019b). Water ingestion as well as skin absorption remain the primary sources of trace elements intake in the milieu (Dhiman and Keshari, 2006; Zhai *et al.*, 2017; Morufu and Clinton, 2017; Raimi and Sabinus, 2017b; Olalekan *et al.*, 2018b; Morufu *et al.*, 2021a). While fluorine remains the 13^th^ most prevalent element in the earth’s crust, it is essential to human life. Elemental fluorine almost never occurs in nature, but fluoride is a shared element that is broadly dispersed in the earth’s crust as well as appears in fluoride form in a number of fluoride rich minerals, like fluoroapatite (Ca_5_(PO_4_)3F), fluorite (CaF_2_), fluorspar, apatite, amphiboles, hornblende, villiaumite (NaF), micas, cryolite (Na_3_AlF_6_), as well as topaz (A_l2_(SiO_4_)F_2_). Heavy groundwater fluoride concentrations are a pervasive problem around the globe; particularly in the global south, where individuals remain disproportionately impacted by fluorosis due to high reliance on groundwater with fluoride contents above the normal threshold. Also, excessive level of fluoride in drinking water might result in a decrease in total erythrocyte, haematocrit value, haemoglobin percentage, as well as protein content. Fluoride (F^−^) is a common groundwater contaminant in loess environments. F^−^ occurs naturally in groundwater as a result of fluoride-containing minerals dissolution. In trace amounts, fluoride is advantageous to the human health as well as can minimize dental caries risk even though encouraging strong bones formation (Li *et al.*, 2014b; Morufu *et al.*, 2021a). According to the WHO (2017), the maximum permissible F^−^ limit in drinking water is 1.5mg/L. Usually, F^−^ in groundwater in the research area is mostly caused by the fluoride-containing rocks dissolution as well as high-fluorine coal (Sun *et al.*, 2014; Morufu *et al.*, 2021a). In this study, [F^−^] in groundwater either fall below or within the WHO/SON/NAFDAC limit. The lowest as well as highest values (0.00 and 0.85mg/L) for raining season (0.00 and 1.02mg/L) for dry season were observed in Ebocha-Obrikom area of Rivers State. Aside from the increased hydrodynamics during gas flaring production, which accelerates F^−^ release from fluoride-containing minerals as well as high-fluorine coals, hydraulic connectivity as well as leakages during gas flaring production might result water mixing from diverse aquifers, affecting concentrations of F^−^ in location 4, 5, 6, 7, and 8 [both raining and dry season]. Chronic exposure toward fluoride raised concentrations in drinking water exceed World Health Organisation recommendation limit of 1.5 mg/L frequently leads to endemic diseases like skeletal as well as dental fluorosis (WHO, 2011). Aluminium had highest recorded value of (0.03) (0.02)mg/L at location 7 during the raining and dry season. No values recorded at location 1, 2 & 9 (for raining season) and location 1, 2, 3 & 9 (for dry season). For these locations, it shows that aluminium does not pose any health and environmental threat to consumption of ground water sources. However, its presence in other locations could be attributed to gas flaring and this of course calls for serious concern. Sodium [Na^+^] is abundant in rocks and soils. It is always present in natural water. It is used medicinally as a laxative (USEPA, 2015; Morufu *et al.*, 2021a). Sodium [Na^+^] varied from 14.33mg/L in raining season to 11.39 mg/L in dry season, which was below the maximum allowable limit of 200 mg/L for drinking uses. Still, the highest [Na^+^], 16.39 mg/L was observed in location 7 of the unconfined groundwater, while the lowest mean [Na^+^], 12.22mg/L, was observed in location 3 of the unconfined groundwater, suggesting that Ebocha-Obrikom oil and gas area of groundwater is affected by more complex factors. Overall, Na^+^ had the highest concentration in location 2, 4, 5, 6, 7 & 8. These ions in groundwater are largely regulated by weathering and water–rock interactions. Also, it can be attributed to longer residence time, mineral composition, and greater water–rock interaction intensity in the aquifer (Olalekan *et al.*, 2021). Thus, the excess of Na^+^ also indirectly indicates the process of ion exchange in water formation (Zhang *et al.*, 2018). In the Ebocha-Obrikom oil and gas area, principal lithology is occupied by crystalline rocks. Therefore, weathering of these rock forming minerals might likely be the chief source for elevated Na^+^ concentration in Ebocha-Obrikom oil and gas area. For potassium (K^+^), it usually exists at low concentrations in groundwater because of weak mobility (Wu *et al.*, 2019b). In this study, [K^+^] ranged from 2.93 mg/L for raining season to 2.82 mg/L for dry season in the aquifer. The highest [K^+^] (3.29 mg/L) was observed in location 8 of the shallow confined groundwater, while the control groundwater had the lowest mean [K^+^], 2.42mg/L at location 9. Thus, K**^+^** is present in a low amount ranging from 2.42 to 3.29mg/L. The concentrations of Ca^2+^, Mg^2+^, and Na^+^ range between (50.31 - 59.98)(40-37 - 50.73), (129.26 -146.67)(149.44 - 184.68), and (12.22 - 16.39)(9.22 - 13.33)mg/L, respectively. Mg^+^ possesses the highest SD value, indicating a very high spatial variability. Thus, He and Wu (2019) reported that the K^+^ in groundwater is one of the very necessary trace elements to maintain human health. K^+^ occurs naturally in drinking water at quantities far below those considered harmful to human health and is the most important nutrient for humans and too much of it might cause constipation (WHO, 2017). However, it is shown that the high level of Sodium (Na^+^) and Potassium (K^+^) in drinking water (beyond the standard limit) may cause hypertension, high blood pressure, hyperkalemia, and often cause a heart attack. For calcium, this element is the most important and abundant in human body and adequate intake is essential for normal growth and health. There is some evidence to show that the incidence of heart disease is reduced in areas served by public water supply with a high degree of hardness, the primary constituent of which is calcium, so that the presence of the element in a water supply is beneficial to health (USEPA, 2015). Hence, calcium is one of the dominant cations in the Ebocha-Obrikom oil and gas area of Rivers State groundwater, ranging from (50.31 – 59.98) (40.37 – 50.73) mg/L with a mean of (54.80 – 45.37) mg/L. According to WHO (2017), the most desirable concentration of Ca^2+^ in groundwater for drinking uses is 75 mg/L and the maximum allowable limit is 200 mg/L. However, the present study results of Ca^2+^ indicated that groundwater samples were below the most desirable limit of 75 mg/L, which can be used for drinking purposes. While, Ca and Mg are present as simple ions Ca^2+^ and Mg^2+^ with the Ca levels varying from tens to hundreds of mg/L and the Mg concentrations varying from units of tens of mg/L (Morufu *et al.*, 2021a). While, calcium and magnesium are essential for the human body (WHO, 2011). They contribute to the formation and solidification of bones and teeth and play a role in the decrease of neuromuscular excitability, myocardial system, heart, and muscle contractility, intracellular information, transmission, and blood contractility (Adimalla and Qian, 2021). They also play a major role in the metabolism of almost all cells of the body and interacts with many nutrients (Olalekan *et al.*, 2021; Morufu *et al.*, 2021a).

However, inadequate, or excess intake of either nutrient can result in adverse health consequences (WHO, 2011). According to WHO (2011) “Inadequate intakes of calcium have been associated with increased risks of osteoporosis, nephrolithiasis (kidney stones), colorectal cancer, hypertension and stroke, coronary artery disease, insulin resistance and obesity. Most of these disorders have treatments, but not cures. Owing to a lack of compelling evidence for the role of calcium as a contributory element in relation to these diseases, estimates of calcium requirement have been made based on bone health outcomes, with the goal of optimizing bone mineral density. To a great extent, individuals are protected from excess intakes of calcium by a tightly regulated intestinal absorption and elimination mechanism through the action of 1,25- dihydroxyvitamin D, the hormonally active form of vitamin D. When calcium is absorbed more than need, the excess is excreted by the kidney in healthy people who do not have renal impairment” (WHO, 2011). In the Ebocha-Obrikom oil and gas producing area of River State groundwater, concentration of magnesium (Mg^2+^) ranged between (129.26 – 146.67) (149.44 – 184.68) mg/L both raining and dry season, with an average mean of (135.18 – 174.61) mg/L (Table 3&4). The major source of Mg^2+^ in the groundwater was magnesium bearing minerals in the host rocks and also animal, domestic, and industrial wastes (Olalekan *et al.*, 2018b; Adimalla and Venkatayogi, 2018; Olalekan *et al.*, 2020a; Marghade *et al.*, 2020; Morufu *et al.*, 2021a). However, all collected groundwater samples were above the maximum allowable limit of 150 mg/L (WHO, 2017). Thus, the high concentration of Mg^2+^ in groundwater may be because of the occurrence of exchangeable Na^+^ in the soil (Henry *et al.*, 2019a; Olalekan *et al.*, 2021). High magnesium hazard (MH) values reveal damage to the soil structure due to a subsequent increase in soil alkalinity. Although, magnesium is significantly less abundant than calcium in rocks and in most natural waters. In addition, magnesium concentrations are much lower in the water than calcium. They are generally less than 50 mg/L, although values higher or equal to 100 mg/L are stored particularly in cold climates (Egbueri and Mgbenu, 2020; Adimalla and Qian, 2021; Morufu *et al.*, 2021a). Magnesium being the fourth most abundant cation in the body and the second most abundant cation in intracellular fluid (WHO, 2017). In the cardiovascular system, magnesium is the candidate element. It plays an important role as a cofactor and activator of more than 300 enzymatic reactions including glycolysis, ATP metabolism, transport of elements such as Na, K and Ca through membranes, synthesis of proteins and nucleic acids, neuromuscular excitability and muscle contraction (Edokpayi *et al.*, 2018; Ukah *et al.*, 2019; Olalekan *et al.*, 2020a; Egbueri and Mgbenu, 2020; Morufu *et al.*, 2021a). That can have hand in various mechanism where the main is the calcium antagonist effect which can be direct or indirect (Olalekan *et al.*, 2018b; Egbueri and Mgbenu, 2020; Morufu *et al.*, 2021a). Low magnesium levels are associated with endothelial dysfunction, increased vascular reactions, elevated circulating levels of C-reactive protein (a proinflammatory marker that is a risk factor for coronary heart disease) and decreased insulin sensitivity. Low magnesium status has been implicated in hypertension, coronary heart disease, type 2 diabetes mellitus and metabolic syndrome. Magnesium deficiency has been implicated in the pathogenesis of hypertension, with some epidemiological and experimental studies demonstrating a negative correlation between blood pressure and serum magnesium levels. However, data from clinical studies have been less convincing (WHO, 2011). Thus, Ca^2+^ and Mg^2+^ are very essential for humans, but excessive intake of them may cause negative human health impact. While, Iron is a lustrous, ductile, malleable, silver-gray metal. Its presence in human tissue for extended periods may cause conjunctivitis, choroiditis and retinitis. A common problem for human is iron deficiency, which may lead to anemia. A man needs an average daily intake of 7mg of iron and a woman 11mg. Presence of Iron in water can lead to change of colour of groundwater (Olalekan *et al.*, 2018b; Olalekan *et al.*, 2020a; Afolabi and Raimi, 2021; Morufu *et al.*, 2021a). In addition, Iron is a highly redox-sensitive element that has low background concentrations in unconfined aquifers, where oxygen is present, but background concentrations of iron can be increased significantly across redox boundaries. It is evident that trace metal can be toxic to human health if they are consumed in excess and accumulated in human bodies (He *et al.*, 2018; Morufu and Clinton, 2017; Raimi and Sabinus, 2017b; Olalekan *et al.*, 2018b; Olalekan *et al.*, 2020a; Afolabi and Raimi, 2021; Morufu *et al.*, 2021a). In this study, the concentration of Fe ranges from (1.21 – 5.16)(0.95 – 4.42) mg/L both raining and dry season, and four (4) samples in raining seasons and two (2) samples in dry season have the Fe concentration higher than the permissible limit for drinking purpose. Thus, indicating fast and pronounced reductive dissolution of iron species in anoxic groundwater. It is well-known that water-quality thresholds may be frequently breached for iron, which occur in groundwater by natural processes, such as the geochemical conditions existing in the aquifer or due to the specific geology of the area. Thus, high concentrations of iron could result in hemochromatosis which is characterized by tiredness, pains in the joints and abdomen (Jonah *et al.*, 2015; Morufu and Clinton, 2017; Raimi and Sabinus, 2017b; Olalekan *et al.*, 2018b; Olalekan *et al.*, 2020a; Afolabi and Raimi, 2021; Morufu *et al.*, 2021a). Zinc is an important mineral apparent by the human today as being excellent biologic and human health significance, mainly concerning prenatal and perinatal growth. Zinc deficiency effects around two billion public in the rising global and is related with several illnesses. In children its sources development delay late sexual development, infection vulnerability, and diarrhea. While, the highest value for zinc was observed at location 3 (0.77)mg/L for raining season and location 4 (1.01)mg/L for dry season. It was noticed that the maximum permissible limit of 3.00mg/L for zinc was not exceeded by any of the locations. Zinc at these limit does not pose serious health and environmental effects though significant values were noticed at locations stated above between the seasons. Thus, zinc could be deposited in those locations due to oil related activities, especially during dry season. Studies have documented that the harmful effects of zinc on people include nausea, lack of moisture, tiredness, pains in the abdomen, lack of coordination of the muscles, and kidney failure. Although a normal dose of zinc is essential to prevent zinc deficiency (Morufu and Clinton, 2017; Raimi and Sabinus, 2017b; Olalekan *et al.*, 2018b; Olalekan *et al.*, 2020a; Afolabi and Raimi, 2021; Morufu *et al.*, 2021a), protracted dosages of zinc could trigger the development of deformed blood cells and the impairment of the pancreas (Ezekwe *et al.*, 2012; Morufu and Clinton, 2017; Raimi and Sabinus, 2017b; Olalekan *et al.*, 2018b; Ukak *et al.*, 2019; Olalekan *et al.*, 2020a; Afolabi and Raimi, 2021; Morufu *et al.*, 2021a). In addition, zinc insufficiency is generally due to unsatisfactory dietary consumption, but can be correlated with malabsorption, Acrodermatitis enteropathica, liver damage, renal damage, sickle cell damage, diabetes, malignancy, and other chronic diseases assemblies at hazard for zinc deficiency contain the elderly, children in rising nations, and individuals with renal deficiency. Signs of mild zinc insufficiency are varied.

Thus, medical consequences contain depressed development, diarrhea, weakness and late sexual development, alopecia, eye and skin abrasions, decreased appetite, changed perception, decreased host protection possessions, defects in carbohydrate utilization, and reproductive spermatogenesis (Morufu and Clinton, 2017; Raimi and Sabinus, 2017b; Olalekan *et al.*, 2018b; Olalekan *et al.*, 2020a; Afolabi and Raimi, 2021; Morufu *et al.*, 2021a). Manganese is a redox-sensitive element, it is influenced by the same processes that determine the reductive dissolution of Fe oxy-hydroxides and as desorption from their surfaces. The maximums value of Mn and Zn are (0.04 – 0.08) and (0.77 – 1.01) mg/L for raining and dry season respectively, and all groundwater samples are suitable for drinking in terms of the Zn concentration, but two (2) samples are not suitable for drinking (location 2&3 – 4&5) because of high Mn concentration in groundwater. While, manganese is an essential nutrient, manganese is neurotoxic at high levels of exposure and evidence suggests that infants could be uniquely vulnerable to its effects (Erikson *et al.*, 2007; Morufu and Clinton, 2017; Raimi and Sabinus, 2017b; Olalekan *et al.*, 2018b; Blanc, 2018; Olalekan *et al.*, 2020a; Afolabi and Raimi, 2021; Morufu *et al.*, 2021a). Manganese exposures in drinking water have been associated with neurodevelopmental outcomes that include reduced IQ or poorer memory, inattention, hyperactivity, impulsivity and motor function in children (Erikson *et al.*, 2007; Oulhote *et al.*, 2014; Morufu and Clinton, 2017; Raimi and Sabinus, 2017b; Olalekan *et al.*, 2018b; Ukah *et al.*, 2019; Olalekan *et al.*, 2020a; Afolabi and Raimi, 2021; Morufu *et al.*, 2021a). Thus, water that exceed the state’s reference dose (RfD) for manganese, which is the highest daily intake that is estimated is likely to cause harmful effects over a lifetime of exposure. This finding should be seen as a wake-up call. As this is not just an Ebocha-Obrikom issue. High water manganese levels are likely prevalent in many parts of the core Niger Delta Region. The study showed that groundwater in Ebocha-Obrikom area of Rivers State contain widely varying amounts of manganese. This study is significant because it calls attention not only to high levels of manganese in groundwater, but also the important fact that many small communities in the Niger Delta region could have elevated manganese in their ground water systems, Thus, residents of the Ebocha-Obrikon community should be aware of the manganese levels in their ground water particularly if they have infants or young children. Cadmium (Cd) are known to increase the risks of lung cancer and renal carcinoma. The highest value for cadmium was observed at location 2, 7 &8 (0.02)mg/L during the raining season and location 4 &7 (0.06)mg/L during the dry season. All values recorded in this study area were above the maximum permissible limit of 0.003mg/l for WHO/SON/NAFDAC. Thus, Cadmium (Cd) is known to cause damage to the kidney, bones in both young and old, also responsible for bronchitis, anaemia (Morufu and Clinton, 2017; Raimi and Sabinus, 2017b; Olalekan *et al.*, 2018b; Olalekan *et al.*, 2020a; Afolabi and Raimi, 2021; Morufu *et al.*, 2021a). Lead is classified as a prevalent toxic metal and a major environmental health hazard, when it enters into the chain through drinking water and crop irrigation. It can accumulate in bone, muscle, liver, kidney and brain. Excessive lead causes problems in the synthesis of hemoglobin, kidney disease, mental retardation, anemia and acute or chronic damage to the nervous system. Pb^2+^ is very toxic to human beings when present in high amounts. Since Pb^2+^ is not biodegradable, once soil has become contaminated, it remains a long-term source of Pb^2+^ exposure. The primary cause of lead’s toxicity is its interference with a variety of enzymes since it binds to sulfhydryl groups found in many enzymes. Lead also interferes in the activity of an essential enzyme called delta-aminolevulinic acid dehydrates, or ALAD and ferrochelatase which are important in the biosynthesis of heme, the cofactor found in hemoglobin. Approximately, contact to lead is growing above period. Extreme level of lead absorptions in the human body can cause death or perpetual harm to the brain, central nervous system and kidneys (USGAO, 2000; Morufu and Clinton, 2017; Raimi and Sabinus, 2017b; Olalekan *et al.*, 2018b; Olalekan *et al.*, 2020a; Afolabi and Raimi, 2021; Morufu *et al.*, 2021a). Lead being one of the most poisonous heavy metals because of its impact on the kidneys and central nervous system (Olalekan *et al.*, 2018b; Olalekan *et al.*, 2020a; Afolabi and Raimi, 2021; Morufu *et al.*, 2021a). The highest value for lead was observed at location 7 (0.14)mg/L during the raining season and location 3 (0.03)mg/L during the dry season. All values recorded in this study area were either within or above the maximum permissible limit of 0.01mg/l for WHO/SON/NAFDAC. Thus, could be said to be of environmental and health concern. As long-term exposure to lead (Pb) can be harmful to the circulatory and central nervous systems. Thus, lead is a hazardous component; it is injurious even in minor quantities. Lead component comes in the human body majorly found in water and food. It can be gasped in powder form of lead in paints, or excess gases from leaded petroleum products. It is originated in minor quantities in several water bodies and food, particularly fish, which remain seriously focus to industrialized toxic waste. Lead reduces enzymes nonfunctional by compulsory to their sulfhydryl group’s additional contribution to a damage in oxidative balance (Morufu and Clinton, 2017; Raimi and Sabinus, 2017b; Olalekan *et al.*, 2018b; Olalekan *et al.*, 2020a; Khan *et al.*, 2020; Afolabi and Raimi, 2021; Morufu *et al.*, 2021a). The capability of lead to permit above the barrier blood and brain is mostly due to its capability to extra for calcium ions. Major toxicity of lead causing the brain prefrontal hippocampus, cerebellum and cerebral cortex can lead to a variability of neurological disorder, such as brain injury, psychological delay, behavior difficulties, nerve injury, and probably Alzheimer’s disease, Parkinson’s disease and schizophrenia (Ezekwe *et al.*, 2012; Morufu and Clinton, 2017; Raimi and Sabinus, 2017b; Olalekan *et al.*, 2018b; Olalekan *et al.*, 2020a; Afolabi and Raimi, 2021; Morufu *et al.*, 2021a). Copper is a reddish metal with a face-centered cubic crystalline structure (Morufu *et al.*, 2021a). It can be found in many kinds of food, in drinking water and in air (Raimi *et al.*, 2018b; Oluwaseun *et al.*, 2019; Raimi *et al.*, 2020a; Raimi *et al.*, 2021b). Long-term exposure to copper can cause irritation of the nose, mouth and eyes and it causes headaches, stomachaches, dizziness, vomiting and diarrhea. According to the Agency for Toxic Substances and Disease Registry (2015), intentionally high uptakes of copper may cause liver and kidney damage even death. Thus, copper is a ductile metal with very high thermal and electrical conductivity. Copper occurs in nature in its metallic form and in ores and minerals. The metal and its alloys have been used for thousands of years. Copper is an essential dietary element for all living organisms in trace amounts, because it plays a major role as a key constituent of the respiratory enzyme complex cytochrome oxidase. Copper had its highest of (0.05)mg/L at location 6 for raining season and (2.81)mg/L at locations 4 during the dry season. All values were below the maximum permissible limit of copper. However, studies have reported that high values of copper could lead to the development of chronic anemia (Morufu and Clinton, 2017; Raimi and Sabinus, 2017b; Olalekan *et al.*, 2018b; Iqbal *et al.*, 2018; Olalekan *et al.*, 2020a; Afolabi and Raimi, 2021; Morufu *et al.*, 2021a). Contamination of drinking water by copper could be by either directly polluting water sources or through rusting of copper pipes and materials. Thus, copper being one of the most common pollutants found in industrial effluents, its extreme consumption of copper leads to gastrointestinal problems, kidney damage, anemia and lung cancer. Copper is lethal for human in the range from 4 to 400 mg/kg of body weight. Lower doses of copper ions can cause symptoms typical of food poisoning (headache, nausea, vomiting, diarrhea) (Morufu and Clinton, 2017; Raimi and Sabinus, 2017b; Olalekan *et al.*, 2018b; Olalekan *et al.*, 2020a; Afolabi and Raimi, 2021; Morufu *et al.*, 2021a). In humans, the liver is the primary organ of copper-induced toxicity. Finally, copper toxicity causes Wilson’s disease in humans. Chromium is a naturally arising metal existing on the geological, by states corrosion ranging from chromium (Utom *et al.*, 2013; Ezekwe *et al.*, 2012; Egbueri, 2020; Adeyeye *et al.*, 2021). Chromium arrives into several environmental mediums (water, soil and air) from an extensive variability of anthropogenic and natural sources with the major discharge upcoming from industry establishing. Industry with the major involvement to discharge chromium contain metal dispensation, tannery services, chromate manufacture, welding stainless steel, and ferrochrome and chrome pigment manufacture. Thus, chromium is a highly toxic element due to its ability to penetrate cell membranes and at high level of exposure can damage the liver and kidney, skin ulceration and also affect the central nervous system (Wang *et al.*, 2016; Kiran *et al.*, 2016; Morufu and Clinton, 2017; Raimi and Sabinus, 2017b; Olalekan *et al.*, 2018b; Olalekan *et al.*, 2020a; Afolabi and Raimi, 2021; Morufu *et al.*, 2021a). In addition, chromium (III) and chromium (VI) have very different toxicity characteristics. Chromium is essential for human nutrition and is considered non-toxic. Levels more than 0.05 mg/L of chromium (VI) in drinking water may cause diarrhoea, vomiting, abdominal pain, indigestion, convulsions, and liver and kidney damage (Morufu and Clinton, 2017; Raimi and Sabinus, 2017b; Olalekan *et al.*, 2018b; Olalekan *et al.*, 2020a; Afolabi and Raimi, 2021; Morufu *et al.*, 2021a). Hexavalent chromium compounds are genotoxic carcinogens. Chronic inhalation of hexavalent chromium compounds increases the risk of lung cancer. Ingestion of chromium (VI) can also cause irritation or ulcers in the stomach and intestines. In this study, chromium had its highest of (1.29)mg/L at location 5 for raining season and (2.81)mg/L at locations 4 during the dry season. All values were above the maximum permissible limit of chromium. Although, chromium does not pose any known serious environmental and public health threat, but it should be closely monitored because the present concentration could be due to gas flaring. The contamination of ground water by chromium could be as a result of exposure to wastes from oil and gas processing facilities which are indiscriminately disposed into open dumps. Chromium in its hexavalent form has been associated with the injurious effects of chromium to mankind. Harmful effects of chromium include liver necrosis and membrane ulcers (Olalekan *et al.*, 2018b; Olalekan *et al.*, 2020a; Morufu *et al.*, 2021a). Hence chromium (Cr) is an essential trace element involved in basal metabolism, too much in the body may lead to negative effects, partly because Cr is a known human carcinogen and anaphylactogen. Environmental exposure of chromium is involving mixtures compound known to produce multi organ poisonousness such as asthma, allergy, renal damage and major effect cancer of the respirational tract in people (Goyer, 2010; Olalekan *et al.*, 2018b; Olalekan *et al.*, 2020a; Morufu *et al.*, 2021a). The major health difficulties seen in human subsequent consumption of chromium mixtures are impatience and ulcers in the small intestine and abdominal, anemia, damage male reproductive system and sperm damage. Nearly persons are tremendously sensitive to chromium, allergic responses containing of severe swelling and redness of the skin have remained noted. A growth in abdominal tumors was observed in people and animals showing to chromium in water. The consumption of excessive quantity was taken of chromium complexes by humans takes resulted in plain respiratory, cardiovascular, gastrointestinal, hematological, hepatic, renal, and neurological effects as part of the sequelae important to death or in patients who persisted as per medical treatment. Though the indication of carcinogenicity of chromium in people and global living thing appears tough (Chen *et al.*, 2009; Morufu and Clinton, 2017; Raimi and Sabinus, 2017b; Olalekan *et al.*, 2018b; Olalekan *et al.*, 2020a; Afolabi and Raimi, 2021; Morufu *et al.*, 2021a). The sulphate (SO_4_ ) concentrations were within permissible limit in the groundwater samples. As shown in Table 3&4, SO_4_ ranged from 0.87mg/L in raining season to 1.01mg/L in dry season. The highest concentrations of (SO_4_ ) were observed in location 4 (0.99mg/L) and the lowest was observed at location 7 (0.84mg/L), these groundwater concentrations of these anions, especially SO_4_ are regulated by hydrogeological and climatic factors as well as anthropogenic factors in groundwater. In particular, evaporation (a climatic factor) is very intense in the area. In addition, hydrogeological (weathering and water–rock interactions) and anthropogenic factors (agricultural and industrial activities) also contributed to increasing major anion concentrations. Major ions determine the general characteristics of groundwater in a given area (Morufu and Clinton, 2017; Raimi and Sabinus, 2017b; Olalekan *et al.*, 2018b; Olalekan *et al.*, 2020a; Afolabi and Raimi, 2021; Morufu *et al.*, 2021a). While significantly lower values could be attributed to unconfined and semi-confined aquifers within water bodies. The highest value of sulphate at location 4 & 2 could be noted for in the water body (Table 3&4), which indicates a high sensitivity of sulphate to changes of geochemical conditions within the aquifer system. Also, it could be stated that gas flaring must have increased the concentration of sulphate during the dry season as against the low levels during the raining season. In addition, it could be attributed to agricultural contamination from fertilizers which later seeped underground to mix with ground water. It could be stated that sulphate is very unstable in the atmosphere from where they are converted into forms suitable for their stay in groundwater’s. It is well known that high concentrations of sulphate may be triggered by dissolution of minerals that control its natural abundance in water or by various land use. The EU Groundwater Directive (2006/118/EC) specifically states sulphate as indicator of the saline intrusion resulting from human activities. An increase of the sulphate concentration due to geogenic processes and human impact is apparent in unconfined aquifers within water body, which can be associated with pronounced human impact from household diffuse pressure or water abstraction in the body. Thus, excessive sulfate use can have a laxative impact. For ammonia, Nutrients salts (ammonia, nitrite and nitrate) are essential to the metabolism and growth of aquatic organisms when its concentration increased; it’s affected the biological equilibrium. In aquatic ecosystem nutrients salts have greatly increased with time through a human activity which causes a problem with water quality (Olalekan *et al.*, 2020a; Raimi *et al.*, 2019a). In some areas, extensive use of mineral fertilizers has led to atmospheric pollution, greenhouse gas emissions (e.g., N_2_O, very important for climate regulation), water eutrophication, and human health risks (Raimi *et al.*, 2018b; Raimi *et al.*, 2020a; Raimi *et al.*, 2021b; Morufu *et al.*, 2021a) thereby negatively affecting the regulating services of soil, air, and water quality (Smith *et al.*, 2013; Morufu and Clinton, 2017; Raimi and Sabinus, 2017b; Raimi *et al.*, 2018b; Olalekan *et al.*, 2018b; Raimi *et al.*, 2020a; Olalekan *et al.*, 2020a; Raimi *et al.*, 2021b; Afolabi and Raimi, 2021; Morufu *et al.*, 2021a). Also, excess inputs of nitrogen can lead to eutrophication, which is the accelerated production of organic matter and the potential development of areas of low dissolved oxygen concentration (hypoxia). The consequences of hypoxia can include the death of benthic organisms, fish kills, reduced growth and disruption of life cycles. Other symptoms of eutrophication include, increases in turbidity, loss of submerged aquatic vegetation and changes in the food web (Okoyen *et al.*, 2020; Olalekan *et al.*, 2021). Thus, ammonia (NH_3_ ) values range from highest at location 6 (2.80) mg/L for raining season to location 3 (4.39) mg/L (Table 3&4). The maximum value is set at 3.0 mg/L according to WHO/SON/NAFDAC standards of potability. For phosphorus, it occurs widely in nature, in plant, in micro-organisms, in animal waste and so on. The significance of phosphorus is principally in regard to the phenomenon of eutrophication of lakes and, to a lesser extent, rivers. Phosphorus gaining access to such water bodies, along with nitrogen as nitrate, promote the growth of algae and other plants leading to blooms, littoral slimes, diurnal dissolved oxygen variation of great magnitude and other related problems. Phosphorus exists in orthophosphate, polyphosphate, organic phosphate and so on (USEPA, 2015). High phosphate levels in drinking water may cause digestive problems in humans and animals. Thus, phosphate had its highest value at location (2 & 3) (0.38) mg/L during the raining season and location 8 (0.55) mg/L during the dry season. All values recorded in this study were above the maximum permissible limit specified by WHO/SON/NAFDAC. This could be due to the fact that farmers in the study area usually nourished the farms with fertilizers of which phosphate fertilizer is one. Also, the high phosphate amount could be attributed to oil and gas related activities and contaminated rains as a results of gas flaring, falling and being retained within ground water within the study area. Also, for Nitrogen, it is an essential element in the biogeochemical cycle; it can be seen as various gases along within organic matter and dissolved species. Naturally, the oxidation state of nitrogen diverges from +5 in NO_3_ ^−^, to − 3 in NH_4_ . It is driven in the following reduction series:

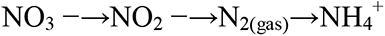

Nowadays, nitrate-contaminated groundwater has become jeopardy in all countries (Raimi *et al.*, 2021a; Olalekan *et al.*, 2021; Morufu *et al.*, 2021a). The uncontrolled fertilizer usage and huge generation of waste with subsequent nitrogen leaching from the soil have gathered serious concerns over the future of nitrogen in aquifers (Wu *et al.*, 2020; Raimi *et al.*, 2021a; Olalekan *et al.*, 2021). Thus, an elevated nitrate concentration in drinking water causes a potentially fatal syndrome where oxygen transport in the bloodstream can be impaired (methemoglobinaemia). While, NO_3_ is usually taken as an indicator of agricultural nonpoint source pollution (Wu and Sun, 2016) and can pose a health risk to humans (Li *et al.*, 2016e). The permissible limit of NO_3_ ^−^ for drinking is 50 mg/L (WHO, 2017). In the groundwater, NO_3_ was in the range of 1.79mg/L to 3.30 [both raining and dry season]. Nitrite pollution within the groundwater is considered to have originated from both industrial and agriculture field. High [NO_3_ ^−^] in drinking water can induce human health hazards, with the occurrence of blue baby and or methaenoglobinaemia patients (Wu and Sun 2016; Morufu *et al.*, 2021a). Typically, it is rare to detect NO_3_ in deep groundwater. Therefore, if NO_3_ is observed in the deep groundwater, it is most likely because of high hydraulic connectivity between the borehole and well aquifers. However, other factors also affect [NO_3_ ] in deep aquifers. Nitrogen associated with the gas flaring could be a source of NO_3_ . Burning of gas flaring could also be a possible source. While, Nitrates (NO_2_ ) is one of the qualitative parameters of groundwater and its enrichment leads to human health implications, hence it entails precise periodic extent. Nitrate is frequently detected in high concentrations in unconfined alluvial aquifers, while its origin, in some cases, can be linked with the existence of elevated chloride and sulphate concentration, particularly in urban areas. While, nitrate (NO_2_ ) is one of the most prevalent contaminants in the groundwater all over the world. Mostly, its high concentration causes immense health disorders, such as methemoglobinemia, gastric cancer, goiter, birth malformations, and hypertension (Zhang *et al.*, 2018; He *et al.*, 2018). NO_2_ are common contaminants in groundwater in the loess areas. However, high NO_2_ concentration in groundwater is regarded as a result of anthropogenic activities such as the application of fertilizer in agriculture and the leaching of sewage wastes or poultry manure. In this study, NO_2_ ranges from (1.61 – 2.33)(2.31 – 4.57) mg/ L, indicating that groundwater samples at location 2, 3, 6 & 9 for raining season are suitable for drinking according to the WHO/SON/NAFDAC drinking water guidelines (WHO, 2017). Thus, availability of nitrogen fixing bacteria on leguminous root nodules that penetrate atmospheric nitrogen into the soil could account for the very low levels of nitrates. However, the concentrations of NO_2_ during the dry season in the studied groundwater samples are high, with the highest value being 4.57 mg/L. This could be due to the twin factor of oil spillage and gas flaring residual effects. Although NO_2_ concentration is below the permissible limit of NO_2_ for drinking purpose (WHO, 2017). As mentioned earlier, oil and gas related activities and agriculture is the main occupation and also the principal income source in the Ebocha-Obrikom oil and gas producing area of River State; therefore, high concentration of nitrate in groundwater is mainly attributed to sewage leaks, household discharges not connected to sewage systems and agricultural activities, where the use of N-fertilizer (synthetic nitrogenous fertilizers and organic manure) is common. In the globe, this result was contrary to the study found in Nuevo-Leon, north-eastern Mexico, where 82% of the sampling locations were influenced by the anthropogenic sources in an agricultural region (Ledesma *et al.*, 2014). A number of studies have proven that a high correlation can be found between agriculture and nitrate concentrations in groundwater (Adimalla and Venkatayogi, 2018; Adimalla *et al.*, 2018a; Zhang *et al.*, 2018; Morufu *et al.*, 2021a; Adimalla and Qian, 2021). Thus, indigenous Ebocha-Obrikom drinking water supply in the 21^st^ century surely faces challenges from algal blooms, lead pipes, nutrient pollution, and other threats. Conclusively, intensive agricultural activities such as fertilizers application and other human activities such as domestic sewage discharge may be the major factors leading to groundwater nitrate contamination. Nitrate pollution is also very common in similar loess areas, because the land and climate conditions are poor in the loess areas, and a large amount of fertilizer has been used to increase the crop yield (Su *et al.*, 2017b; Zhai *et al.*, 2017; Raimi *et al.*, 2020b; Isah *et al.*, 2020b; Morufu, 2021; Hussain *et al.*, 2021a). For Nickel, it is usually found in human tissues and, under situations of high exposure, these levels may rise significantly. In the universal population, contributions to the people intake of nickel from drinking water are normally less significant than dietary intake and absorption is the most significant way of exposure. The consumption of nickel is dependent on its physicochemical procedure, with water-soluble methods (chloride, nitrate, sulphate) present further readily consumption. Thus, the values for nickel was higher at location 4 (1.00)mg/L for raining season and location 3 (1.40)mg/L for dry season respectively. The values were higher than the WHO/SON/NAFDAC tolerable limits of 0.02 mg/L. The nickel values differed remarkably. Even though nickel has been identified as a vital trace metal, it could also be highly poisonous at higher doses. Hair loss, lung fibrosis, allergies of the skin, eczema, and various degrees of kidney and heart poisoning have been associated with humans exposed to high concentrations (Ezekwe *et al.*, 2012; Utom *et al.*, 2013; Obiadi *et al.*, 2016; Morufu *et al.*, 2021a). Nickel also has the propensity of replacing iron and zinc in the body, thus interfering in the normal biochemistry (Olalekan *et al.*, 2018b; Olalekan *et al.*, 2020a; Morufu *et al.*, 2021a). While differences in distribution happen as a role of way of exposure, the solubility of the nickel element and period after exposure, the major target organs for nickel-induced general toxicity are the lungs and the higher respiratory tract for inhalation exposure and the kidney for oral exposure. Additional object organs contain the cardiovascular system, the immune system and blood. Human exposure to extremely nickel-polluted in water has the probable to produce a range of pathological affects. Amongst them are skin allergies, lung fibrosis, cancer of the respiratory tract and iatrogenic nickel toxicity. Nickel exposure has been conveyed to produce hematological effects in both humans and animals. Although no reproductive effects have been linked with nickel exposure to humans. In addition, the geographic distributions of TPH are depicted in Table 3 & 4, respectively. The research areas of location 1, 2, 3, 4 & 6 have higher TPH concentrations, while location 5, 7 & 8 have lower TPH concentration and in location 9 TPH was not detected for raining season. The content of TPH in groundwater, on the other hand indicated that locations 2, 3, 4, & 7 had higher concentration above WHO/SON/NAFDAC standards, while location 1, 5, 6 & 8 had concentration below permissible standards. But location 9 did not show any presence of TPH for dry season. The findings revealed that five (5) locations in the raining season and four (4) locations in the dry season did not fulfill the WHO/SON/NAFDAC criteria. Accordingly, the result demonstrates that TPH concentrations in drinking water remain significantly greater, implying that water quality may have an unfavorable effect on fish survival, eggs and production of larvae as well as ecosystem maturation. Because of the relatively high tidal velocities, the pollution is spread out over a vast area. The river turbulence, on the other hand, meant that the spill was broken down into smaller, less harmful pieces. There is also concern about the lengthy period required for total biodegradation of the heavier components, which contain very hazardous aromatic compounds with low boiling points. The high saturated boiling point as well as aromatic hydrocarbons possess the potential to interfere with aquatic creatures’ responses toward chemical stimuli, like sex desire with equally severe repercussions. The utmost harmful pollution aspect, and definitely all pollutions, is the toxicant amplification problem, because several of the crude oil components remain chemically stable as well as not easily metabolized or eliminated once taken, making them available in the food chain at all times. The high TPH values in those sites are a cautionary sign that everything is not well, since some water company and vendors use ground water for production as well as sell it in places nearby or as far away as Yenagoa and Imo. Aside from its deadly effects, oil can induce death via producing narcosis, which causes animals to get detached from substrate. oil coating can induce death through asphyxiation in other creatures. Oil coatings on the water surface in damaged areas impede light transmission and thus photosynthetic primary production. In the interim, the physical as well as chemical consequence untoward effects as well as oil effects remain long-lasting. As a result, we must not forget that the general pollution effect of this sort and magnitude occurrence on water bodies as well as ecosystem is significantly more problematic to anticipate. As a result, total recovery may perhaps take close to 20 years. In summary, heavy metals causing neurological impairments, anemia, kidney failure, immunosuppression, gastrointestinal and respiratory irritation, abnormalities of skeletal system, inflammation of liver, cancer of liver, cardiovascular diseases after chronic exposure. Diagrammatically, the main contaminants effects on human health (see figure 7 below) is represented thus:

**Figure 7.**
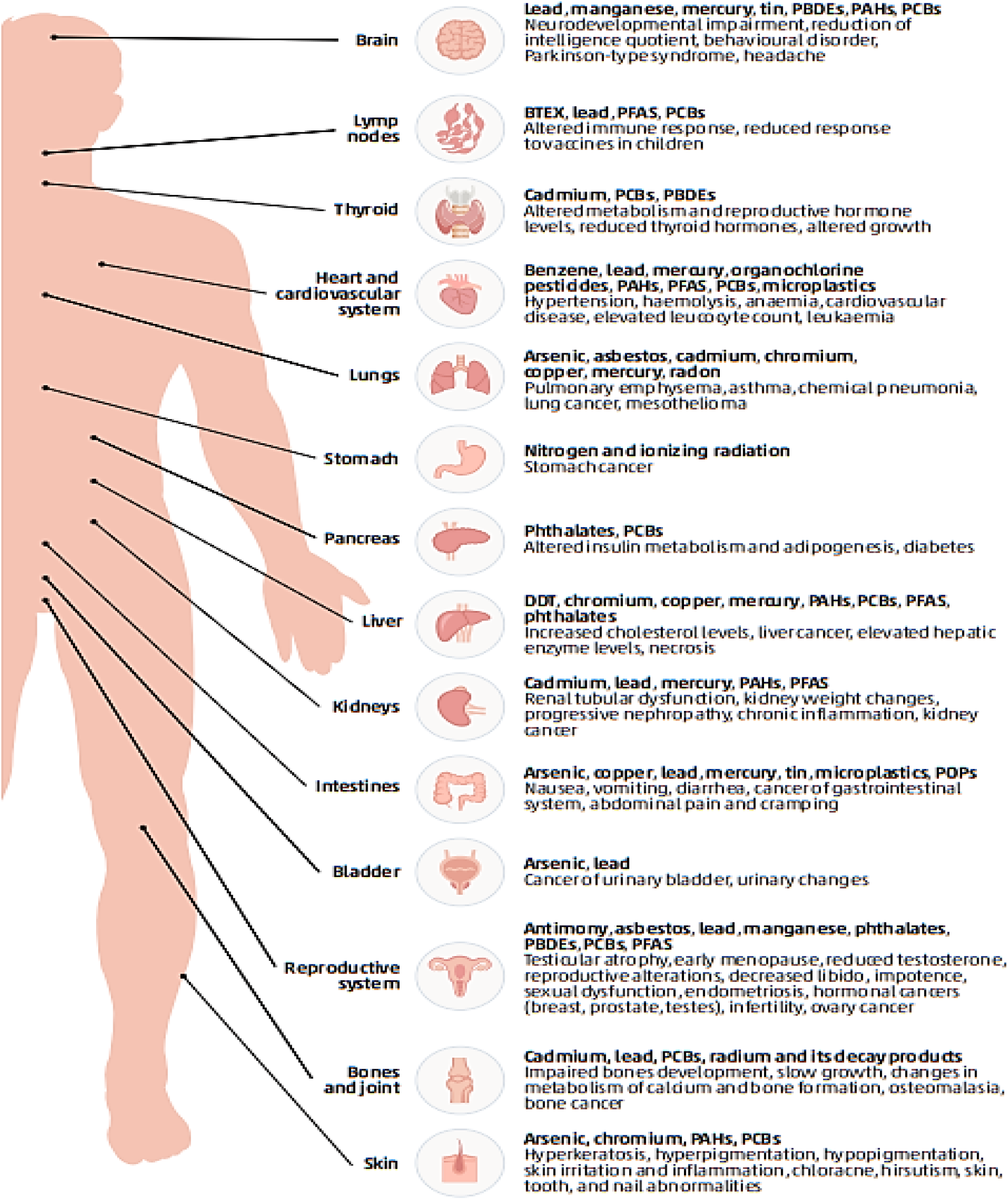
Main effects of contaminants on human health, indicating the organs or systems affected and the contaminants causing them. **Source:** Adapted from Morufu *et al.*, (2021a)

## 7. Conclusion and Recommendations

Pollution is a worldwide issue with no boundaries. Contaminants can be found on every continent, even in the most distant locations, and they can easily be moved from one country to the next. Thus, since ancient times, human activities have produced thousands of different manmade chemical compounds and naturally occurring components with potential toxicity into the environment. These contaminants have a long residence duration in the environment, ranging from hundreds to thousands of years, and are found all over the world. Hence, groundwater pollution has become as one of the most serious threats to human health, but its consequences extend far beyond the dimensions as well as contaminants, as it can have irreversible consequences for human as well as ecosystem health, result in severe economic losses as well as social inequities and jeopardize the 2030 Agenda for Sustainable Development achievement. Hence, preferred groundwater is the water resource supply around the world, particularly in the global south, where water is scarce and of poor water quality. the available assumption as well as sustainable confined groundwater is hydrochemistry, which must remain concerned preferentially. The Ebocha-Obrikom oil and gas producing area of River State, was chosen as the study area in order to gain awareness into the hydrochemistry as well as perspective of groundwater health and to make available decision-useful information as well as assist in taken action to solve the urgent threats facing societies across the Niger Delta, while fostering understanding as well as collaboration through a diverse stakeholder’s range of actionable solutions. The following highlights are offered after the broad research findings:

❖ Development partners as well as local governments must be involved in the artificial recharge schemes implementation as well as maintenance at the community level.
❖ The relevant stakeholders have an urgent task ahead in closing down open wells in the Niger Delta region of Nigeria, for the sake of population likely to be affected through them, since they live near gas flaring area and make use of polluted groundwater, also because the indigenous population breath in toxins released from gas flaring. Unless we act, groundwater pollution growth is inevitable in the Niger Delta.
❖ This study has proven that groundwater pollution exposure had a bigger negative influence on indigenous inhabitants’ life expectancy than COVID-19.
❖ In addition, open lines of communication amongst policy makers, academia as well as society remain essential to guarantee that decision makers as well as other stakeholders have access to timely, science-based information on the possible hazards caused via contaminants.
❖ Eventually at the community level, public and school students should be educated on groundwater quality and correct management through a series of seminars, short films, and other activities. Furthermore, seasonal groundwater quality monitoring should remain conducted, as well as any necessary precautions to prevent further groundwater contamination.
❖ To improve groundwater quality, suitable management techniques such as regulating human activities, implementing water treatments, raising public awareness, as well as establishing a groundwater quality monitoring network are advised.
❖ Human interference (indiscriminate disposal of drainage wastes and uncontrolled use of agricultural pesticides) should be more effectively and strictly managed, as this is the most significant method for preventing groundwater contamination.
❖ It is strongly advised that frequent monitoring as well as assessment of overall water resources supplies be promoted. Waste disposal, land usage, and agricultural methods that help to preserve the quality of water resources should all be applied. Before consumption, the water should be thoroughly boiled.
❖ The first step in water pollution management is identifying and assessing risk at potentially polluted sites. If contamination at a given site is at levels that can harm humans, information about that site should be collected at the appropriate governmental level and made public, and remediation or risk-minimization actions should be taken accordingly, especially if the site is used for food production or as a water reservoir for human consumption.
❖ In light of the current global trend scenario of worsening groundwater pollution, stronger political, business, as well as social commitment is required to identify alternatives to the usage of extremely harmful pollutants as well as increased research investment in prevention as well as cleanup.
❖ Enhanced cooperation as well as partnership remain required to enable knowledge availability, the exchange of successful experiences, as well as worldwide access to safe and sustainable technologies, that leave no one behind.
❖ Agip should immediately begin replacing all old pipes in the Ebocha-Obrikom Oil Fields as soon as possible, and should collaborate with other agencies to complete a comprehensive Joint Investigation Visit (JIV) report. Furthermore, fair compensation should be provided to the impacted victims of Agip carelessness because their means of livelihood have been annihilated.

## Data Availability

The data that support the findings of this study are summarized in the various tables and are available without undue reservation from the corresponding author while the data are submitted to the Figshare web portal:
https://doi.org/10.6084/m9.figshare.14273234.v1.

https://doi.org/10.6084/m9.figshare.14273234.v1

## Acknowledgments

I thank Prof. Mynepalli K.C. Sridhar, Prof. Oyeyemi Abisoye S, Prof. Kalada G. Mcfubara, Prof. B.E. Akpan, Prof. I.M. Aprioku as well as all anonymous reviewers, for feedback and discussions that helped to substantially improve this manuscript.

## Funding

The water sampling and analysis was supported by AGIP Research Department.

## Conflict of Interest

The authors declare that the research was conducted in the absence of any commercial or financial relationships that could be construed as a potential conflict of interest.

## Data Availability Statement

The data that support the findings of this study are summarized in the various tables and are available without undue reservation from the corresponding author while the data are submitted to the Figshare web portal: https://doi.org/10.6084/m9.figshare.14273234.v1.

## Supplementary Material

The Supplementary Material for this article can be found online at: Raimi, Morufu (2021): Water Quality Parameters in Gas Flaring Area of Ebocha-Obrikom of Rivers State, Nigeria. Figshare. Dataset.

https://doi.org/10.6084/m9.figshare.14273234.v1. https://figshare.com/s/c7f5d0e75e096a20a8e1.

## Notes

### Competing Interest Statement

The authors have declared no competing interest.

### Funding Statement

This study did not receive any funding

